# The Heterogeneous Effects of Lockdown Policies on Air Pollution^∗^

**DOI:** 10.1101/2023.05.11.23289832

**Authors:** Simon Briole, Augustin Colette, Emmanuelle Lavaine

**Author notes:** We are grateful to Richard Le Goff, Kévin Jean and Sandrine Mathy for their help and comments. We thank INERIS for providing us access to the data used in this work. We also thank the support of the ADEME grant AQACIA 2162D0019 - Air-COV project.

## Abstract

While a sharp decline in air pollution has been documented during early Covid-19 lockdown periods, the stability and homogeneity of this effect are still under debate. Building on pollution data with a very high level of resolution, this paper estimates the impact of lockdown policies on *PM*_2.5_ exposure in France over the whole year 2020. Our analyses highlight a surprising and undocumented increase in exposure to particulate pollution during lockdown periods. This result is observed during both lockdown periods, in early spring and late fall, and is robust to several identification strategies and model specifications. Combining administrative datasets with machine learning techniques, this paper also highlights strong spatial heterogeneity in lockdown effects, especially according to long-term pollution exposure.

**JEL Codes:** C23, I18, Q53

## 1 Introduction

A growing body of evidence highlights a sharp decline in air pollution in many countries during early Covid-19 lockdown periods (Venter et al., 2020; Berman & Ebisu, 2020; Mahato et al., 2020; Dang & Trinh, 2021; Brodeur et al., 2021). This effect appears to be driven by sharp reductions in human mobility and economic activity (Faridi et al., 2021). However, some recent studies tend to mitigate this result and the stability and homogeneity of this effect is still under debate (Adam et al., 2021; Schneider et al., 2022; Bartoňová et al., 2022). In particular, these studies suggest that the effects of lockdown policies are likely be very heterogeneous depending on the geographic area and the type of air pollutant considered. Understanding the heterogeneity in lockdown effects on air pollution and analyzing the mechanisms mediating this heterogeneity is key given the large detrimental effects of air pollution on health and economic outcomes (Äıchi & Husson, 2015; Deryugina et al., 2019).

This paper investigates the effects of lockdown policies on *PM*_2.5_ and *NO*_2_ exposure in France, a country where *PM*_2.5_ emissions primarily originate from the residential and tertiary sector. In such a context, the theoretical impact of lockdown policies on *PM*_2.5_ is unclear. On the one hand, the curtailment of economic activities and reduced human mobility (Dang & Trinh, 2021) during lockdown periods tend to decrease the level of *PM*_2.5_. On the other hand, increases in residential heating or biomass burning (Sicard et al., 2020) as well as increases in *O*_3_ (Adam et al., 2021) may enhance the formation of *PM*_2.5_.

Like in many countries, France was hit by two Covid-19 epidemic waves in 2020, one in early spring and the other in late fall, and implemented lockdown policies during the two corresponding periods. We show that these two periods coincided with strong increases in *PM*_2.5_ concentration, by about 25% with respect to non-lockdown periods. By contrast, nitrogen dioxide (*NO*_2_), which is mainly related to transport emissions, decreased drastically during both periods.

To reach these conclusions, our analyses draw on daily air pollution data available for the whole French territory over the 2015-2020 period. This dataset provides daily measures of *PM*_2.5_ and *NO*_2_ exposure with a very high level of resolution (2×2km grid level). Our estimation strategy builds on the comparison between air pollution during lockdown and non-lockdown periods, controlling for the same baseline difference in previous years. This approach allows us to control for any seasonal and geographic correlations between pollutant emissions, meteorological conditions and air pollution at a very fine scale.

This paper contributes to the growing literature studying the causal impact of lockdown policies on *PM*_2.5_. Causal evidence of lockdown-induced pollution remains scarce and primarily based on the analysis of early phases of the pandemic (Brodeur et al., 2021; Dang & Trinh, 2021). In this paper, we leverage long-term air pollution data to build a measure of *excess* pollution exposure, defined as the deviation in 2020 *PM*_2.5_ pollution exposure with respect to a counterfactual (pre-lockdown) period – namely the average of pollution exposure over the 2015-2019 period. This strategy deals not only with geographic correlations but also with seasonal correlations between pollutant emissions, meteorological conditions and air pollution. On top of this, our analyses build on a massive dataset of nearly 50 millions observations, defined at the 2×2km grid level, a much finer geographic scale than the one used by previous papers which typically build on county-level or national-level analyses.

Our results highlight the importance of documenting the heterogeneity in the effects of lockdown on air pollution. While most of the existing evidence point to a positive effect of lockdown on air quality, we document a sharp increase in *PM*_2.5_ in France during lockdown periods. Building on Machine Learning techniques, we also show that the impact of lockdown is stronger in the areas with the highest levels of long-term exposure to air pollution. These results are consistent with the fact that *PM*_2.5_ emissions in France primarily originate from the residential and tertiary sector as well as with the fact that people spent much more time at home than usual during the two lockdown periods (Brandily et al., 2021). They also suggest that the effects of lockdown policies on air pollution can vary a lot both across and within countries and crucially depends on the type of pollutant considered and its sources of emissions.

The remainder of this paper is organised as follows. Section 2 presents background information on air pollution, lockdown policies and the data exploited. Section 3 provides graphical evidence on the evolution of air pollution in France in 2020. Section 4 presents the estimation strategy and the main results from our regression analyses. Finally, Section 5 concludes with a brief discussion of the main results outlined in this paper.

## 2 Background and data

### 2.1 Air Pollution

Our analyses draw on air pollution data provided by the French National Institute for Industrial Environment and Risks (*INERIS*), available over the 2000-2020 period for the whole of metropolitan France. This gridded reanalysis pollution data combines numerical modeling data from the *CHIMERE* chemistry-transport model (Menut et al., 2013) with measures of air quality from monitoring stations to correct for ground observations. Gridded reanalysis pollution data are computed by *INERIS* in order to build population-wide measures of background pollution exposure.^1^ They provide hourly measures of exposure to the four main regulated pollutants: *PM*_2.5_, *PM*_10_, *NO*_2_, and *O*_3_. These measures are available at a very fine geographical level, namely at the 2km x 2km level. We aggregate pollution data at the daily level, for each of the 134,459 grid cells located in metropolitan France.

Our analyses focus on exposure to *PM*_2.5_ and *NO*_2_, whose negative effects on human health have long been documented in the epidemiological and economic literature.^2^ The thresholds recommended by the *World Health Organization* with respect to *PM*_2.5_ are 5 *µg/m*^3^ for annual average exposure and 15 *µg/m*^3^ for daily average exposure. The thresholds for *NO*_2_ are 10 *µg/m*^3^ for annual exposure and 25 *µg/m*^3^ for daily exposure (WHO, 2021). *PM*_2.5_ exposure in France has slightly but continuously decreased over the last decade, in line with the global trends observed in European countries (Sicard et al., 2021; Salesse, 2022) and in the US (Currie & Walker, 2019; Currie et al., 2023). Despite this encouraging trend, average exposure to *PM*_2.5_ in France in 2020 is 7.8 *µg/m*^3^, which is well above the recommended threshold.^3^ Beyond the annual averages, the daily thresholds set by the *WHO* are very regularly exceeded over the 2015-2020 period, even when considering weekly averages *PM*_2.5_ exposure (Figure A1). Figure A2 also shows that there is a strong spatial heterogeneity in *PM*_2.5_ exposure over the French territory.

More than 55% of *PM*_2.5_ emissions in France originate from the residential and tertiary sector, while 15% originate from the transport sector (Lavaine et al., 2020; Citepa, 2022). By contrast, *NO_x_* emissions primarily originate from the transport sector (58%). For both pollutants, the agricultural and industrial sectors respectively account for 10-17% of emissions. As outlined in Figure A1, which depicts weekly average levels of *PM*_2.5_ exposure in France over the 2015-2020 period, there is a strong seasonality in *PM*_2.5_ concentration, due to both anthropogenic emissions and weather conditions.

### 2.2 Weather Data

On top of pollutant emissions, air quality conditions are also determined by changes in weather conditions.^4^ To account for these factors, we match our pollution data with weather data from the the French national institute for meteorological data monitoring (*Météo France*). We match each grid cell from pollution data to the closest weather station,^5^ and we derive the following daily weather variables for each grid cell: total precipitations, surface maximum temperatures, surface minimum temperatures, wind speed and wind direction. For this last parameter, we build dummies for each of four directions: North (below 45◦ or above 315◦), East (between 45◦ and 135◦), South (between 135◦ and 225◦) and West (between 225◦ and 315◦).

### 2.3 Socio-economic data

To investigate the heterogeneity of our results, we also match our pollution data with the *Filosofi* 2017 dataset, an administrative fiscal dataset produced by the French National Institute for Statistics (*INSEE*). This gridded dataset provides socio-economic indicators at a very local level, namely at the 1×1km grid level, for the whole French population in 2017. For each grid cell, the dataset provides the number of inhabitants and their age distribution, the average standard of living (i.e., the disposable household income divided by the number of consumption units), the share of poor households, the share of owner households and the share of single-parent households. This dataset also includes detailed information on housing conditions, allowing us to compute for each grid cell the the share of households living in houses, the share living in collective dwellings, the share living in subsidized housing, the total surface area of dwellings and the age of houses.

### 2.4 Lockdown policies

#### 2.4.1 Lockdown periods in France in 2020

Like in many European countries, France was hit by two distinct Covid-19 epidemic waves in 2020. These two waves respectively peaked in April, with 15,479 excess deaths observed over this single month, and November, with 12,537 excess deaths (Brandily et al., 2021).^6^ In both cases, the French government reacted by taking extraordinary containment measures. As COVID-19 first spread in the country in early 2020, the government decided of a national lockdown on March 17 and that eventually lasted until May 11. This first lockdown was the most stringent: all workers stayed home except if their activity was deemed essential for the country. A second lockdown was decided on October 30 and continued until December 15. This second lockdown was slightly less stringent than the previous one and got repealed quicker, with the end of a first phase after a month when all shops opened again. Using Google mobility data, Brandily et al. (2021) show that, all over the 2020 year in France, time spent at home appears much higher when a lockdown policy is in place, and slightly more so during the first lockdown than the second.

#### 2.4.2 Oxford Stringency Index

To better account for the variety in the nature and intensity of policy responses taken by the French government over the 2020 year, and to the likely resulting variations in air pollution, we also exploit the Oxford Stringency Index (OSI). This index was developed by Hale et al. (2021) to measure the overall strictness of lockdown restrictions at a given point in time in a given country. It is constructed as the mean of eight policy measures, namely school closing, workplace closing, canceling of public events, restrictions on gathering, closed public transportation, stay-at-home requirements and travel restrictions (both internally and internationally). The OSI is computed as the mean of the standardized policy measures (between 0 and 1), so that each policy measure contributes equally to the index, independently from its number of levels. The OSI is then rescaled to have values between 0 (no response) and 100 (maximum response in every possible policy measure). Figure A3 in the appendix shows the evolution of this index in France over the 2020 year. It can be noted that the period in between the two lockdown periods is not characterized by the absence of restriction measures, confirming the interest of using a more continuous measure of policy restrictions.

## 3 The Effects of Lockdown Policies on Air Pollution: Graphical Evidence

In the remainder of the paper, we explore the effects of lockdown policies on air pollution. Before moving on to our econometric investigations, we start by providing simple graphical evidence on the evolution of exposure to *PM*_2.5_ and *NO*_2_ over the 2020 year. Figure 1 first depicts the evolution of the daily national exposure to *PM*_2.5_ in France in 2020, contrasting lockdown periods (in black) with regular periods (in gray). The figure reveals a marked increase in the national exposure to *PM*_2.5_ during the two lockdown periods. In particular, it shows a clear discontinuity in *PM*_2.5_ exposure, with a jump right after the start of each lockdown period, respectively in March 17 and October 30.^7^ Such discontinuity is less visible *NO*_2_ exposure, and the average levels of exposure don’t seem to be sharply different during lockdown periods, as opposed to regular periods of 2020 (Figure 2).

**Figure 1:**
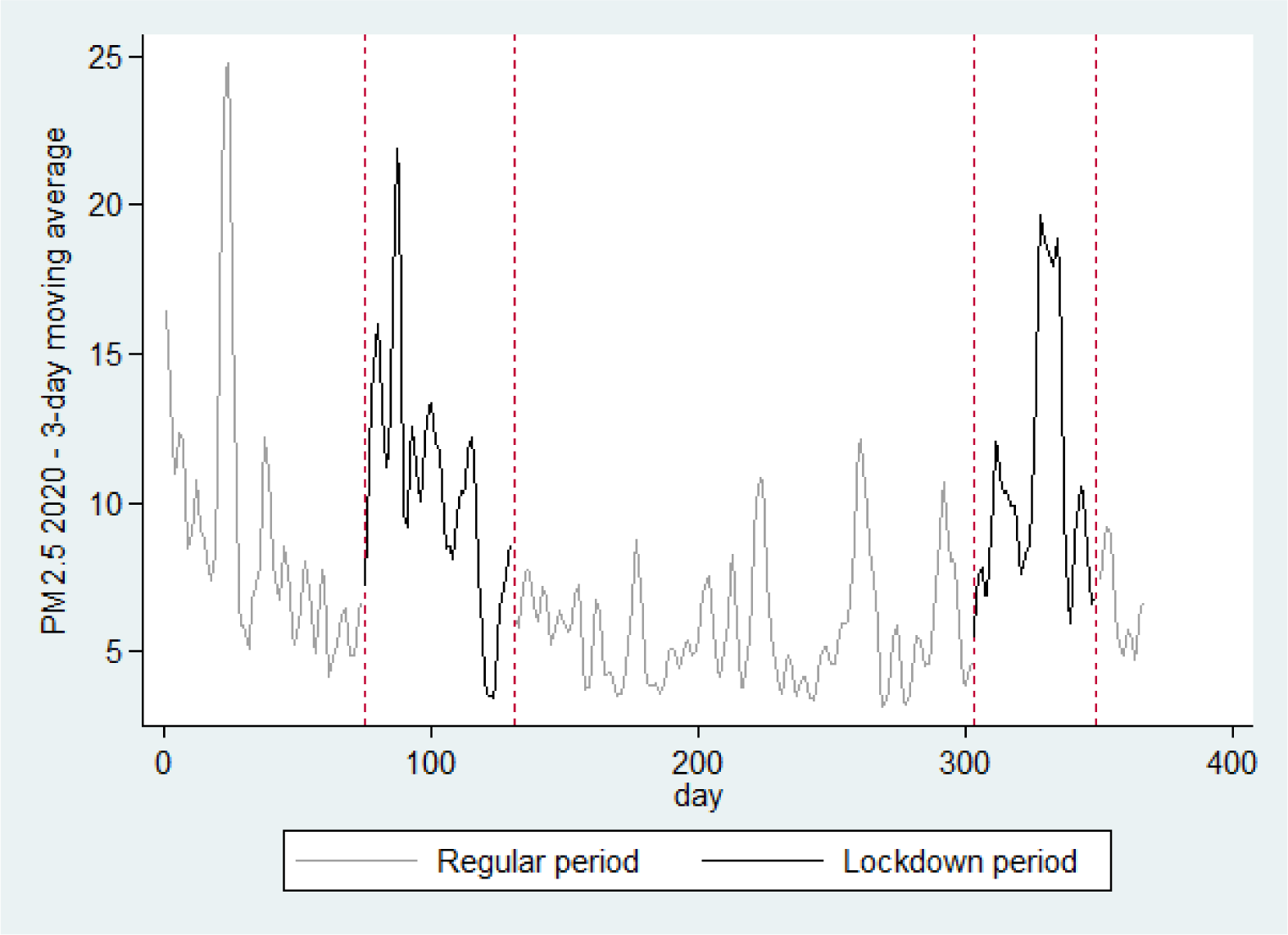
*PM*_2.5_ exposure in France in 2020 Note: This figure plots the 3-day moving average of the daily national exposure to *PM*2.5 in France in 2020, computed at the 2×2km grid level. Each grid is weighted by the number of individuals living in the corresponding area.

**Figure 2:**
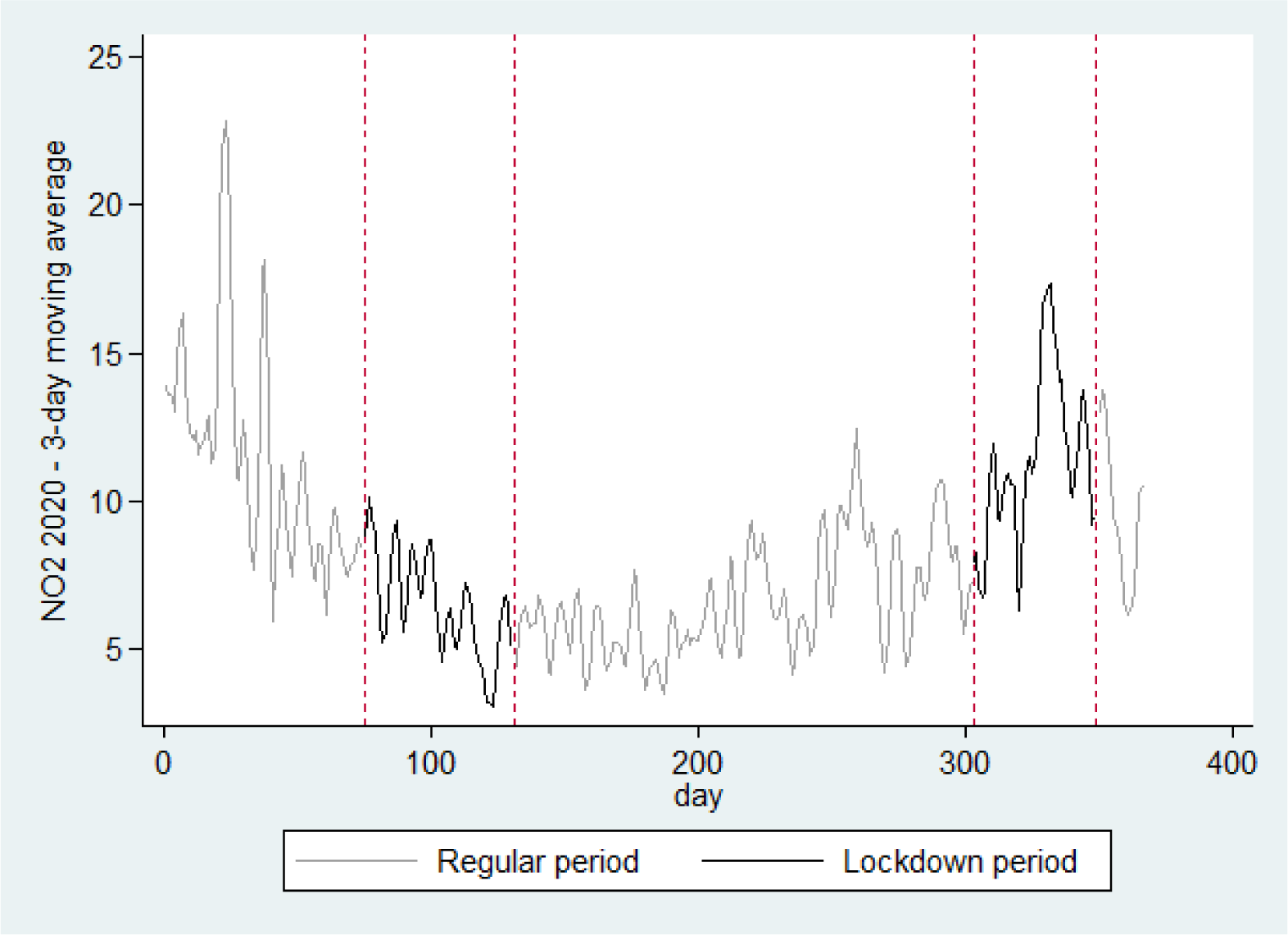
*NO*_2_ exposure in France: 2020 Note: This figure plots the 3-day moving average of the daily national exposure to *NO*2 in France in 2020, computed at the 2×2km grid level. Each grid is weighted by the number of individuals living in the corresponding area.

While Figure 1 suggests a positive effect of lockdown restrictions on *PM*_2.5_ exposure, it could also reflects the strong seasonality in *PM*_2.5_ concentrations highlighted in Figure A1. To take one step further, we compute for each grid cell the difference between the level of exposure to *PM*_2.5_ a given day of the year in 2020 and the average level observed during that same day of the year over the 2015-2019 period. This allows us to account for spatial and temporal regularity in terms of pollutant emissions and weather conditions at a very fine scale (i.e., controlling for day-of-the-year *×* grid fixed-effects).

Figures 3 and 4 replicate Figures 1 and 2, but plotting this outcome expressed in difference instead of the raw level of exposure. These two figures first confirm that both *PM*_2.5_ and *NO*_2_ concentration levels are lower in 2020 than in previous years, a result that have been widely documented in the recent literature (Venter et al., 2020). However, Figure 3 also reveals that lockdown periods systematically coincide with sudden and sustained increases in *PM*_2.5_ *differential* exposure levels, in line with analyses based on raw exposure levels. It seems harder to conclude on the relationship between *NO*_2_ differential exposure levels and lockdown policies from this graphical analysis. The next section presents results from econometric analyses aiming at testing the robustness and exploring the heterogeneity of these effects.

**Figure 3:**
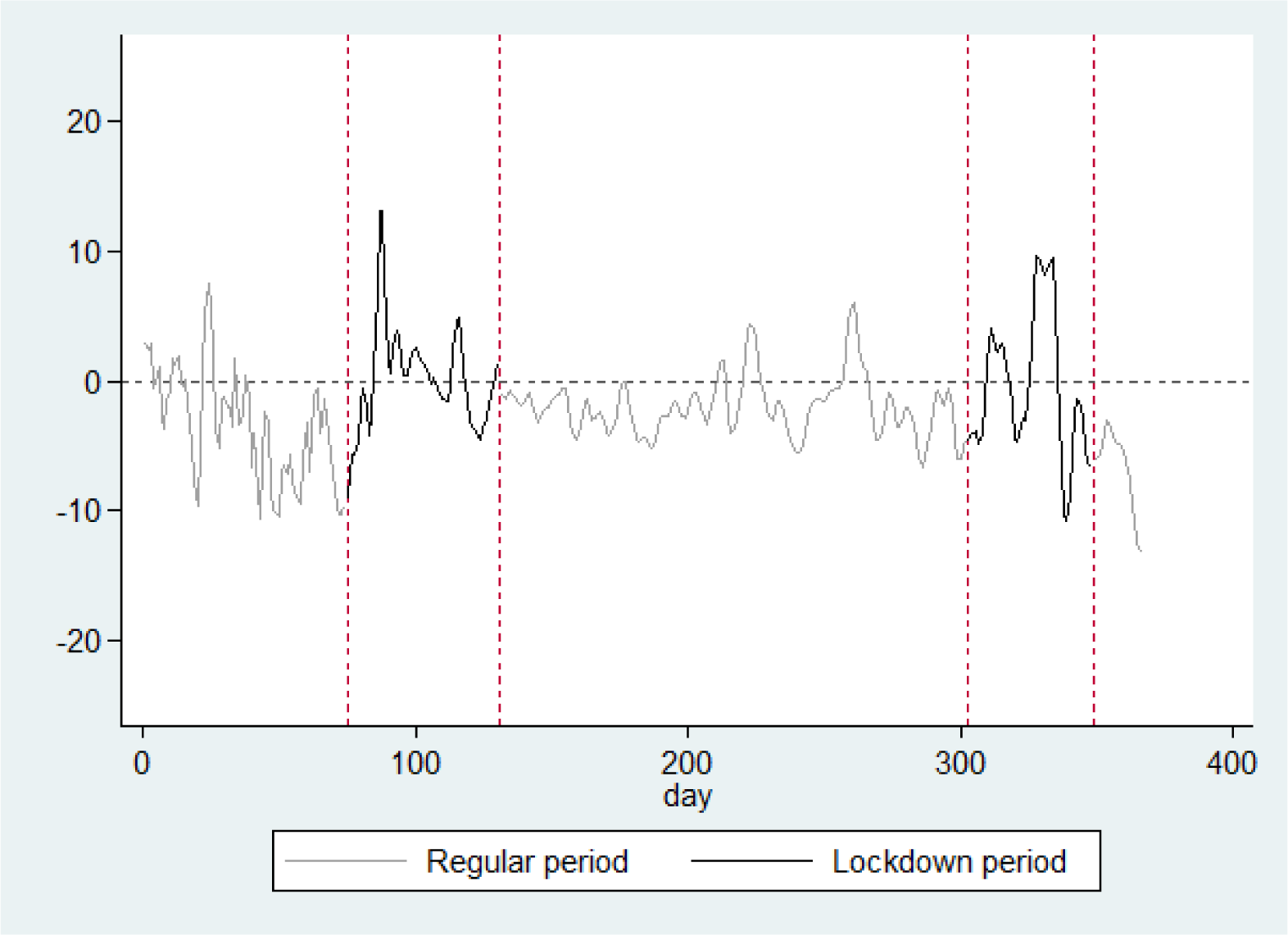
*PM*_2.5_ exposure in France: 2020 vs 2015-2019 Note: This figure plots the 3-day moving average of the difference between daily national exposure to *PM*2.5 in France in 2020 vs 2015-2019, computed at the 2×2km level. Each grid is weighted by the number of individuals living in the corresponding area.

**Figure 4:**
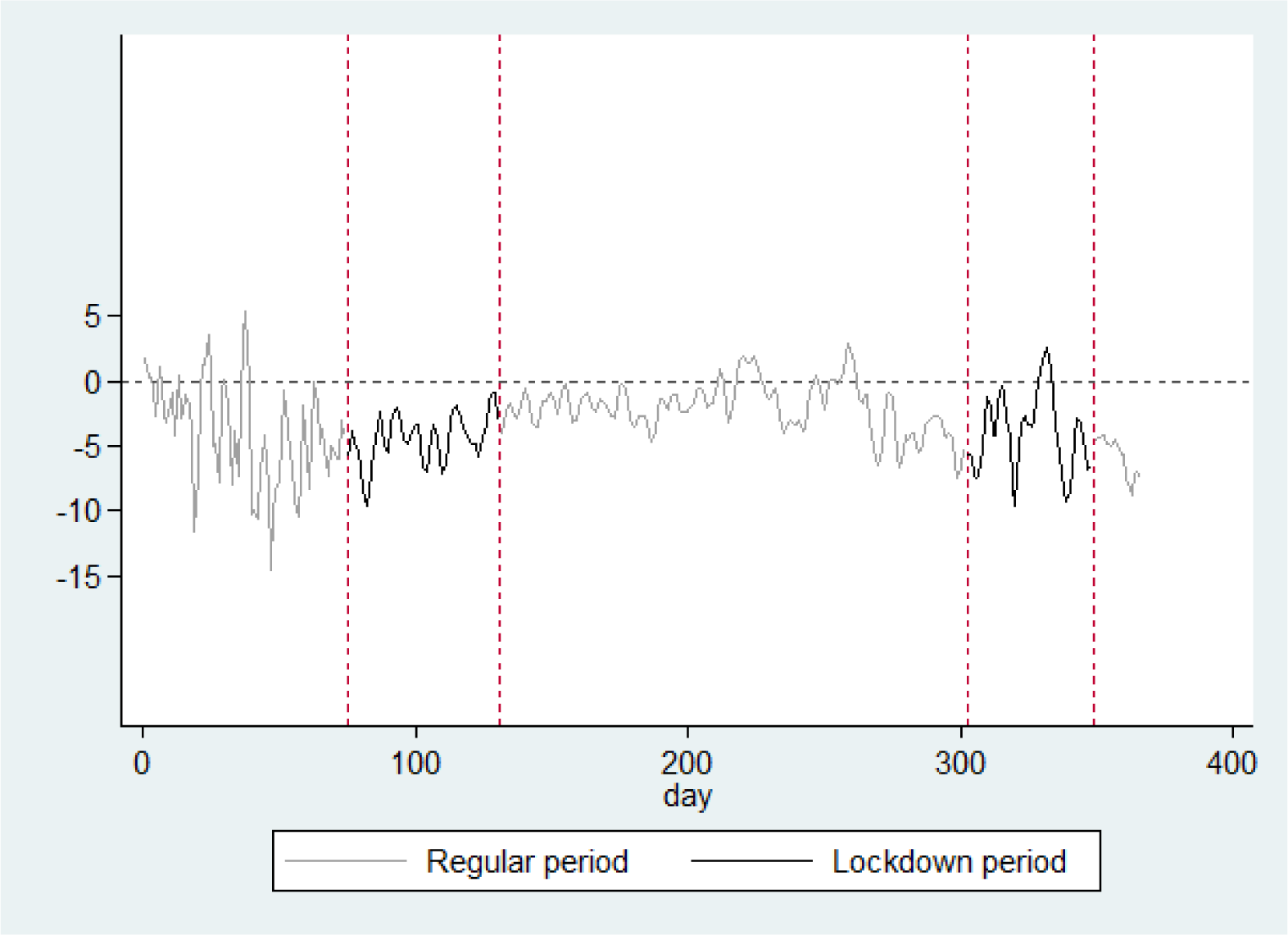
*NO*_2_ exposure in France: 2020 vs 2015-2019 Note:This figure plots the 3-day moving average of the difference between daily national exposure to *NO*2 in France in 2020 vs 2015-2019, computed at the 2×2km level. Each grid is weighted by the number of individuals living in the corresponding area.

## 4 The Effects of Lockdown Policies on Air Pollution: Regression Analysis

### 4.1 Econometric approach

Our objective is to estimate the effects of restrictions imposed by lockdown policies on *PM*_2.5_ exposure. As a baseline approach, we compare the levels of *PM*_2.5_ exposure observed in 2020 in each grid cell during lockdown vs. regular periods, controlling for local weather conditions and grid cell fixed effects. In practice, we estimate the following model:

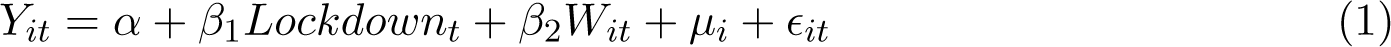

where *Y_it_* represents *PM*_2.5_ exposure in grid *i* on day *t* of year 2020 and *Lockdown_t_* is a dummy indicating lockdown periods. *µ_i_* represents the full set of grid fixed effects. This parameter allows us to control for any geographic differences in *PM*_2.5_ exposure that is constant over time across grid cells. *W_it_*represents the set of weather control variables described in Section 2.2.^8^ We cluster all standard errors at the day level, which is the level at which our “treatment” variable (i.e., the lockdown dummy) varies.

While our baseline approach accounts for the confounding effects of local weather conditions as well as for any geographic differences in *PM*_2.5_ exposure that is constant over time, it could be biased by seasonal effects on pollution emissions or concentrations. In particular, human activities may not be constant throughout the year, with some periods corresponding systematically to an increase in pollutant emissions and others to a reduction in emissions. These seasonal effects may also vary over space, depending on the type of area considered (e.g., rural vs urban, North vs South, etc.). To account for these factors, we estimate a model that allows *PM*_2.5_ exposure to depend on grid-specific *×* day-of-the-year fixed effects (*κ_it_*).^9^ We compute *Y_it_^∗^*, the difference between *PM*_2.5_ exposure in 2020 in grid cell *i* on day-of-the-year *t* and the average *PM*_2.5_ exposure in that same grid cell and day-of-the-year over the 2015-2019 period. We then estimate the following (implicit) difference-in-difference (DiD) model:^10^

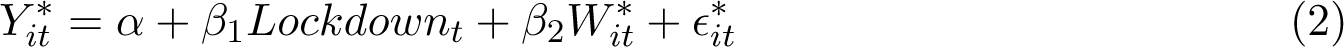

where *Y_it_^∗^ = *Y*_*it*,2020_ − *Ŷ*_*it*,2015−2019_*. This model therefore compares pollution exposure in lockdown vs. regular days in 2020 vs. 2015-2019. It implicitly controls for year, day-of-the-year, grid cell and grid cell *×* day-of-the-year fixed effects. It therefore accounts for the global decreasing trend in *PM*_2.5_ observed over our period of observation. It also accounts for any seasonal correlation between human activities and pollution, allowing this correlation to vary at a very local level (i.e., 2×2km grid cell). The parameter of interest *β*_1_ captures the causal effects of lockdown on *PM*_2.5_ exposure under the assumption that, absent lockdown (and once controlled for local weather shocks), the difference in *PM*_2.5_ between lockdown and regular periods observed in 2020 would have been similar to that same difference in previous years (2015-2019). In Appendix B, we provide extensive evidence that our results are robust to several alternative identification strategies and/or specifications: our main conclusions hold with a model based on a regression discontinuity design (Section B1), with an explicit DiD model (Section B3), when changing weather controls (Section B2) or when relying on randomization-based inference (Section B4).

While our main analyses compare days of lockdown with days of regular periods in a binary way, we also implement regressions using the Oxford Stringency Index. To do so, we estimate Equations (1) and (2) using this index instead of the dummy indicating lockdown periods. This allows us to capture more variability in the restriction measures implemented all over the 2020 year.

### 4.2 Average effects

Table 1 shows the results of the estimation of Equations (1) and (2), using a dummy indicating lockdown periods on the one hand and the Oxford Stringency Index on the other hand. As can be seen in column (1), *PM*_2.5_ exposure levels are higher during lockdown than regular periods in France, once controlled for local weather conditions and grid cell fixed effects. The point estimate is significant at the 1% level and suggests that on average, there was a 2.5 *µg/m*^3^ higher exposure during lockdown periods. This result is confirmed by the estimation of a model that accounts for seasonal effects in pollution emissions and meteorological conditions. As can be seen in column (2), *PM*_2.5_ exposure levels are 1.9 *µg/m*^3^ higher during lockdown than during regular periods of 2020, compared to the same baseline difference in previous years. This lockdown-induced increase in *PM*_2.5_ is consistent with the fact that *PM*_2.5_ emissions in France primarily originate from the residential and tertiary sector as well as with the fact that people spent much more time at home than usual during the two lockdown periods (Brandily et al., 2021). We further investigate the stability of this effect across lockdown periods. Interestingly, results presented in Table 2 show a very high stability in the effects of lockdown policies on *PM*_2.5_ exposure: in both specifications, the effect is virtually the same across the two lockdown periods. This result is important with respect to the external validity of our conclusions. It also further reduces concerns that meteorological factors may drive the estimated effects.

**Table 1:** Effects of lockdown on *PM*_2.5_ exposure in 2020

| | (1)<br>$PM_{2.5}$ 2020 | (2)<br>2020 vs 2015-2019 | (3)<br>2020 | (4)<br>2020 vs 2015-2019 |
| --- | --- | --- | --- | --- |
| Lockdown dummy | 2.5048***<br>(.44992) | 1.9054***<br>(.49143) |  |  |
| Oxford Stringency Index |  |  | .02052**<br>(.00998) | .03905***<br>(.00843) |
| Weather controls | ✓ | ✓ | ✓ | ✓ |
| Observations | 49211994 | 49211994 | 46119437 | 46119437 |

**Table 2:** Effects of lockdown on *PM*_2.5_ exposure in 2020: distinguishing the two lockdown periods

| | (1)<br>$PM_{2.5}$ 2020 | (2)<br>2020 vs 2015-2019 |
| --- | --- | --- |
| First lockdown | 2.5921***<br>(.61298) | 1.8955***<br>(.72495) |
| Second lockdown | 2.4362***<br>(.55303) | 1.9135***<br>(.61200) |
| Observations | 49211994 | 49211994 |

Standard errors in parentheses
\* $p < 0.10$ , \*\* $p < 0.05$ , \*\*\* $p < 0.01$

Analyses based on the Oxford Stringency Index, which accounts for restriction measures in a more flexible way than the simple lockdown dummy, provide very similar results. The estimates presented in columns (3) and (4) both suggest a positive and significant effect of these measures on *PM*_2.5_ exposure, both in level and in difference with respect to the 2015-2019 period. The point estimate in the last column implies that a 45 points increase in the index (i.e., the average difference in the index between lockdown and regular periods) would result in a 1.8 *µg/m*^3^ increase in *PM*_2.5_ differential exposure between lockdown and regular periods, compared to the baseline difference in previous years.

Table 3 replicates the analyses presented in Table 1 with *NO*_2_ as the outcome. The results presented in this table suggest that lockdown restriction measures affect negatively the level of *NO*_2_ exposure. The point estimate presented in column (2) implies that *NO*_2_ exposure levels are 1.9 *µg/m*^3^ lower during lockdown than during regular periods of 2020, compared to the same baseline difference in previous years. This lockdown-induced reduction in *NO*_2_ is consistent with the fact that *NO*_2_ emissions in France primarily originate from the transport sector and with the sharp decline in transport emissions observed in France during lockdown periods due to reduced human mobility. It is also consistent with previous studies on the effects of lockdown on *NO*_2_ exposure (Venter et al., 2020; Berman & Ebisu, 2020; Mahato et al., 2020; Schneider et al., 2022).

**Table 3:** Effects of lockdown on *NO*_2_ exposure in 2020

| | (1)<br>$NO_2$ 2020 | (2)<br>2020 vs 2015-2019 | (3)<br>2020 | (4)<br>2020 vs 2015-2019 |
| --- | --- | --- | --- | --- |
| Lockdown dummy | -1.2756***<br>(.35583) | -1.9966***<br>(.27802) |  |  |
| Oxford Stringency Index |  |  | -.04621***<br>(.0064) | -.01303*<br>(.00677) |
| Weather controls | ✓ | ✓ | ✓ | ✓ |
| Observations | 49211994 | 49211994 | 46119437 | 46119437 |

### 4.3 Heterogeneous effects

The implementation of Covid-19 lockdown policies coincides with an increase in *PM*_2.5_ exposure in France in 2020. An important question, however, is whether this effect have been homogeneous over the territory. In this section, we build on Machine Learning techniques to explore this issue and to shed light on the distribution of treatment effects over the territory according to socio-economic indicators and long-term pollution levels.

#### 4.3.1 Machine Learning analysis

In this section, we take a Machine Learning approach to explore the spatial heterogeneity of lockdown effects in a data-driven manner, allowing us to include a high number of heterogeneity dimensions without being subject to the risk of overfitting. More specifically, we implement the generalized random forest (GRF) procedure introduced by Athey et al. (2019). This approach has been implemented in several recent studies exploring the heterogeneity of treatment effects in a variety of contexts (see e.g., Carter et al. (2019); Allcott et al. (2020); Haaland & Roth (2020); Sylvia et al. (2021); Briole et al. (2022)). In our setting, it makes it possible to predict treatment effects for each grid cell individually using all available information on its characteristics and to test the existence of heterogeneity in lockdown effects.^11^ To train our procedures, we use two measures of long-term pollution exposure defined at the grid level, namely average exposure to *PM*_2.5_ and *NO*_2_ over the 2009-2019 period, as well as all the socio-economic indicators described in Section 2. Denoting *Y* our main outcome of interest, *L* the dummy indicating lockdown periods and *Z* the set of baseline covariates, this procedure starts by growing two regression forests to construct estimates *Y* (*Z*) and *L*(*Z*) of *E*(*Y |Z*) and *E*(*L|Z*). Building on these two estimates, the procedure then grows a causal forest to construct an estimate *S*(*Z*) of the conditional average treatment effect *s*_0_(*Z*) = *E*(*Y*_1_ *− Y*_0_*|Z*), where *Y*_1_ and *Y*_0_ represent grid cells’ potential outcomes in treated and non-treated states. Finally, following Athey & Wager (2019) and Chernozhukov et al. (2018), it is possible to test for the existence of heterogeneity in *s*_0_(*Z*) by regressing *Y − Y* (*Z*) on *C* = *S^̄^*(*L − L*(*Z*)) and *SD* = (*S*(*Z*) *− S^̄^*)(*L − L*(*Z*)), where *S^̄^* represents the average of *S*(*Z*), and by looking at the significance of the regression coefficient of *SD*, which provides an estimate of *Cov*(*S*(*Z*)*, s*_0_(*Z*))*/V ar*(*S*(*Z*)). Let’s denote this regression coefficient by *β*. Rejecting *H*_0_ : *β* = 0 implies rejecting that the actual variance of *s*_0_(*Z*) is zero. It also implies rejecting that the causal forest estimates of treatment effects do not represent relevant predictors of the actual treatment effects.^12^

We conducted this test by considering *Y_it_^∗^*, namely the difference between *PM*_2.5_ exposure in 2020 in a given grid cell on a given day-of-the-year and the average *PM*_2.5_ exposure in that same grid cell and day-of-the-year over the 2015-2019 period, as our main outcome. To implement this test, we randomly selected a subsample representing 5% of the total number of observations in our main sample (i.e., 2,146,677 observations) for computational reasons. The detailed results of this test are given in Table 4. They show that the null hypothesis that *β* = 0 is unambiguously rejected for our main outcome variable, highlighting the presence of a significant spatial heterogeneity in the effects of lockdown on *PM*_2.5_ exposure. Further evidence of this spatial heterogeneity is provided by Figure C1, which depicts the distribution of conditional average treatment effects (CATEs). As the figure reveals, CATEs vary considerably, taking a range of values from 0 to +6.

**Table 4:** Generalized Random Forests: Tests for Heterogeneity

|  | (1)<br>*PM*<sub>2.5</sub>: 2020 vs 2015-19 |
| --- | --- |
| $\beta$ coefficient | 0.35 ***<br>(0.01) |
| Most important variables | Long-term *PM*<sub>2.5</sub><br>Long-term *NO*<sub>2</sub><br>Inhabitants' age<br>Average income<br>Collective dwellings |

Note: This table shows the results of the test for heterogeneity in treatment effect proposed by Chernozhukov et al. \(2018\), which seeks to fit the Conditional Average Treatment Effect (CATE) as a linear function of the out-of-bag causal forest estimates. This test is performed on our main outcome that measures *PM*<sub>2.5</sub> differential exposure in 2020 (as compared to 2015-2019), using a random subsample representing 5% of our main sample. The first row of the table shows the main $\beta$ coefficient of this regression and its standard errors (in parentheses), clustered at the day-of-the-year level. The next 5 rows show the 5 most important variables determining the heterogeneity of lockdown effects. \* $p < 0.10$ , \*\* $p < 0.05$ , \*\*\* $p < 0.01$ .

To further explore the sources of treatment effect heterogeneity, it is possible to identify the variables that are most often used by the causal forest procedure to grow trees and predict individual treatment effects. More specifically, for each heterogeneity variable, it is possible to count the proportion of splits on this variable used by the procedure, giving a higher weight to a split the earlier it occurs in the development of a tree. When we conduct this analysis, we find that the most important sources of treatment heterogeneity relate to long-term pollution exposure (both *PM*_2.5_ and *NO*_2_).^13^

To illustrate the importance of long-term pollution exposure, Figures 5 (a) and 5 (b) respectively plot the distribution of long-term *PM*_2.5_ and *NO*_2_ exposure by deciles of the CATEs distribution. These figures reveal a clear positive and linear relationship between the impact of lockdown on *PM*_2.5_ exposure as predicted by the GRF procedure and long-term pollution exposure in the grid cell, for both *PM*_2.5_ and *NO*_2_ long-term exposure. Analyses based on a classical regression approach confirm the significance of these heterogeneous effects (Tables C1 and C2). These analyses also show that the correlation between lockdown effects and long-term exposure is much stronger when considering *PM*_2.5_ rather than *NO*_2_.

**Figure 5:**
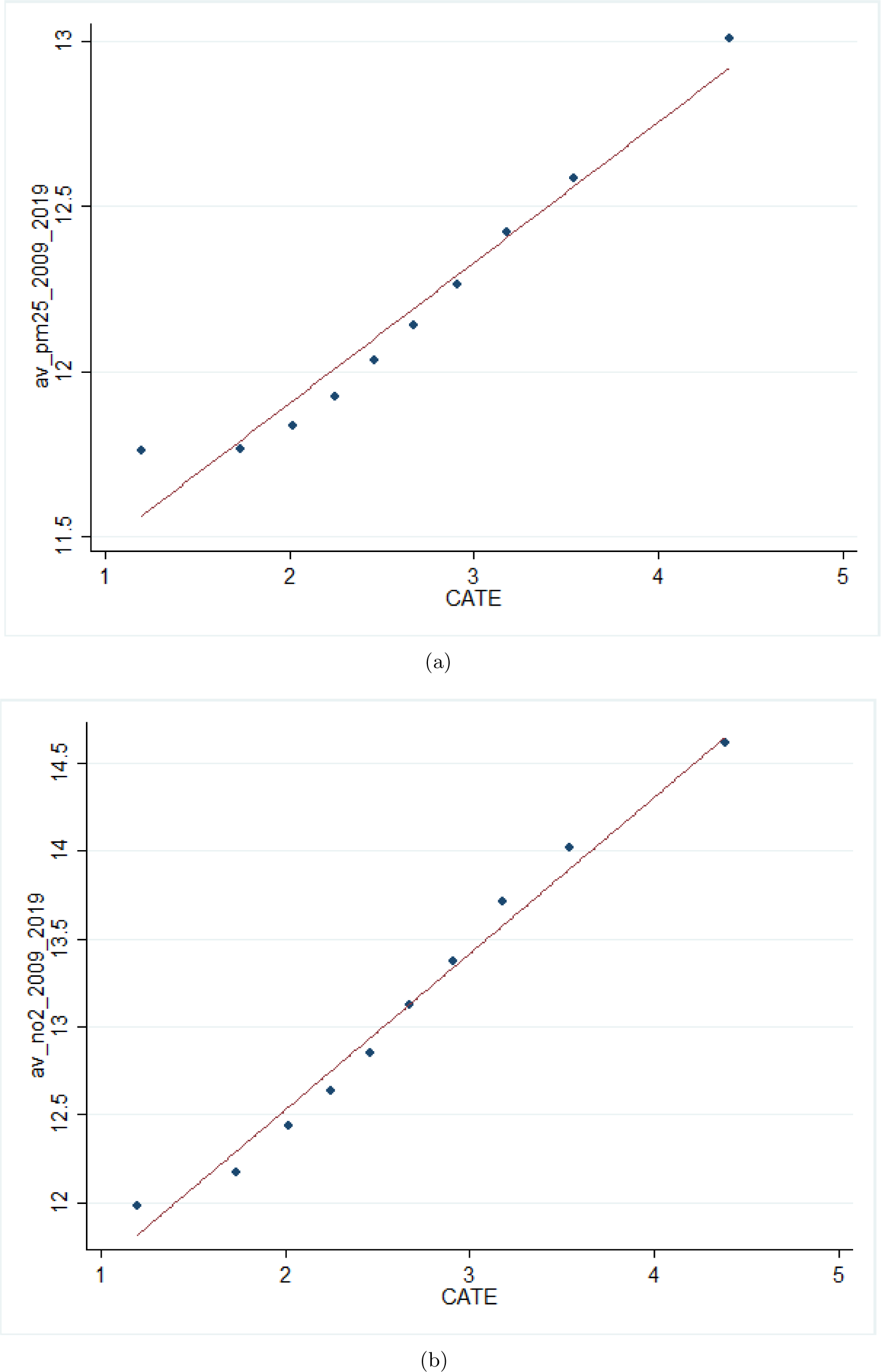
Conditional Average Treatment Effects and Long-term Pollution Exposure *.N..o..t..e..: .*Figure 5 (a) shows the distribution of grid cells average *PM*2.5 exposure over the 2009-2019 period by deciles of the conditional average treatment effects estimated from the GRF procedure. Figure 5 (b) shows the distribution of grid cells average *NO*2 exposure over the 2009-2019 period by deciles of the conditional average treatment effects estimated from the GRF procedure.

Finally, on top of long-term pollution levels, the effects of lockdown on *PM*_2.5_ exposure appear to be heterogeneous along some socio-economic indicators. As illustrated in Figures C2, C3 and C4, the predicted impact of lockdown tends to be stronger in grid cells with younger inhabitants, with a higher share of households living in collective dwellings and with higher household income. Altogether, these results suggest that the positive impact of lockdown on *PM*_2.5_ exposure tend to be stronger in urban than rural settings.

## 5 Discussion and conclusion

While a recent literature highlights the positive effects of lockdown policies on air quality, evidence based on causal methodologies remains scarce and focuses on the early implementation of lockdown policies. This paper builds on daily air pollution data available at a very fine geographical level in France to estimate the impact of lockdown on *PM*_2.5_ and *NO*_2_ exposure. Building on an implicit DiD strategy, we contrast excess pollution during days of lockdown to excess pollution during days of regular periods of 2020, compared to the same baseline difference in previous years. Our results first point to a general decrease in *PM*_2.5_ and *NO*_2_ exposure in 2020 compared to previous years, especially in the most polluted areas. More surprisingly, our analyses also highlight a strong increase in *PM*_2.5_ during both lockdown periods, with exposure levels about 2 *µg/m*^3^ (+25%) higher than during the rest of the year. This result holds when accounting for seasonality in human activities and meteorological conditions in a very flexible way, or when using alternative identification strategies or specifications.

Based on machine learning techniques, we also find strong spatial heterogeneity in this effect, with the increase during lockdown periods being more pronounced in areas with a high level of long-term pollution exposure. This result is consistent with the fact that *PM*_2.5_ emissions primarily originate from the residential and tertiary sectors in France as well as with the fact that people spent much more time at home than usual during lockdown periods. To the extent that long-term exposure is able to capture it, the increase of residential heating in areas with high energy poverty may be another source of explanation for this strong pattern of heterogeneity.

Our results are contrast with previous finding in the literature documenting strong improvement in air quality related to lockdown policies implemented during early phases of the Covid-19 crisis. They highlight the necessity to account for the spatial and temporal heterogeneity of lockdown effects on air pollution, especially with respect to the main sources of pollution. Understanding the impact lockdown policies on *PM*_2.5_ and *NO*_2_ exposure is crucial from a health perspective. An extensive literature documents the detrimental of these pollutants on mortality, hospitalizations related to cardiovascular, respiratory or metabolic problems and associated economic outcomes (Deryugina et al., 2019; Äıchi & Husson, 2015). In addition to usual health outcomes, *PM*_2.5_ exposure could also play an important role in the severity of Covid-19 symptoms (Persico & Johnson, 2021; Isphording & Pestel, 2021). Finally, the oxidative potential of *PM*_2.5_ emitted during lockdown periods could be strong. While assessments of the chronic and acute effects of particulate matter on human health tend to be based on mass concentration, with particle size and composition, a recent literature shed light on the oxidative potential concentration mostly associated with anthropogenic sources, in particular with fine-mode secondary organic aerosols largely from residential biomass burning and coarse-mode metals from vehicular non-exhaust emissions (Daellenbach et al., 2020). Besides, a review about air quality changes during lockdown periods highlights the enhanced formation of secondary *PM*_2.5_ during these periods (Adam et al., 2021). Future research should further explore the causal impact of secondary *PM*_2.5_ exposure on health during lockdown periods.

## Data Availability

All data produced in the present study are available upon reasonable request to the authors

## Main Figures

## Main Tables

## Appendix A Descriptive Statistics

**Figure A1:**
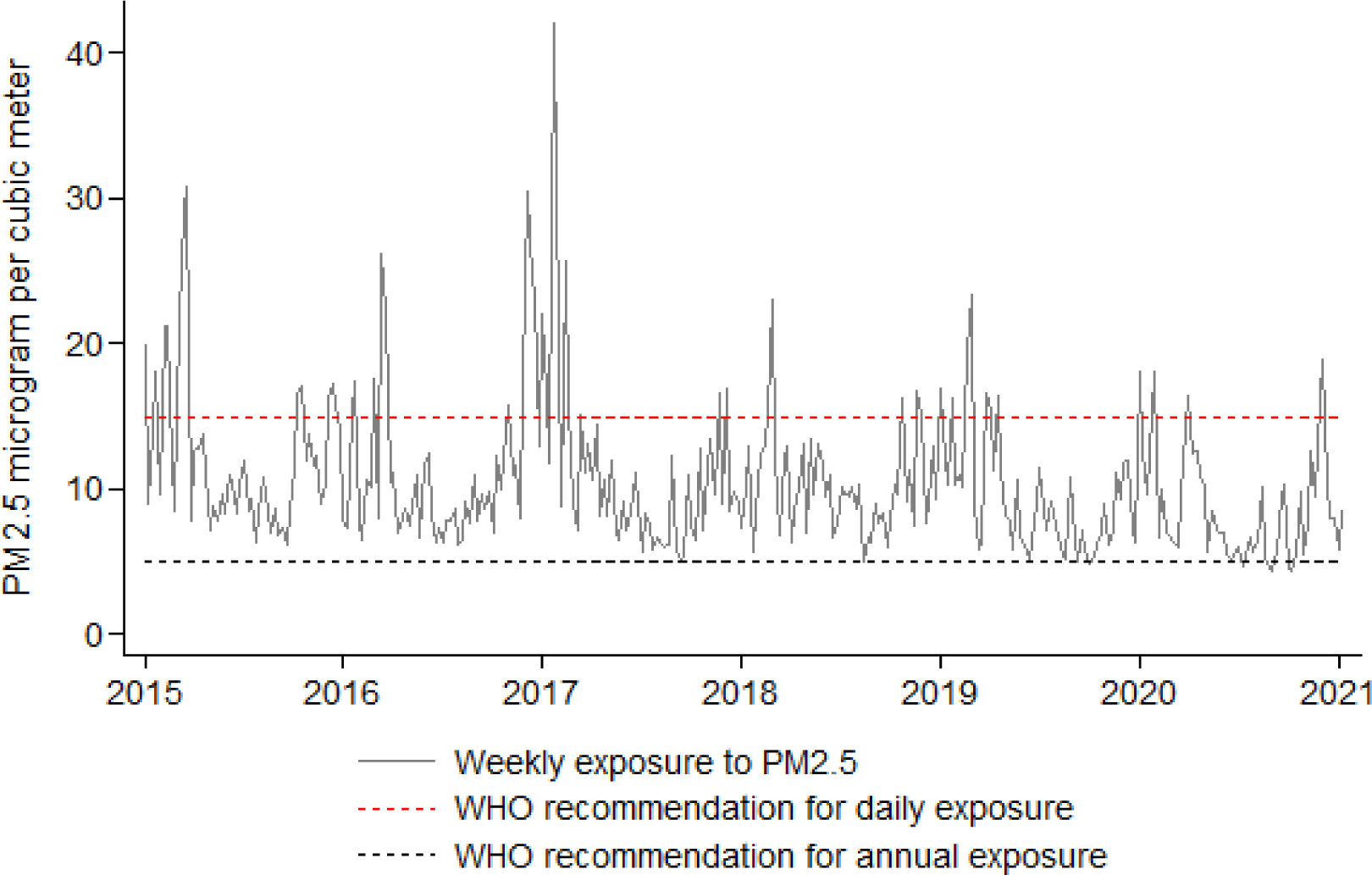
Evolution of *PM*_2.5_ exposure in France (*µg/m*^3^) Note: This figure represents the evolution of the population-weighted weekly average of *PM*2.5 exposure in France over the 2015-2020 period. The World Health Organization recommendations for annual and daily exposure are 5 *µg/m*^3^ and 15 *µg/m*^3^, respectively.

**Figure A2:**
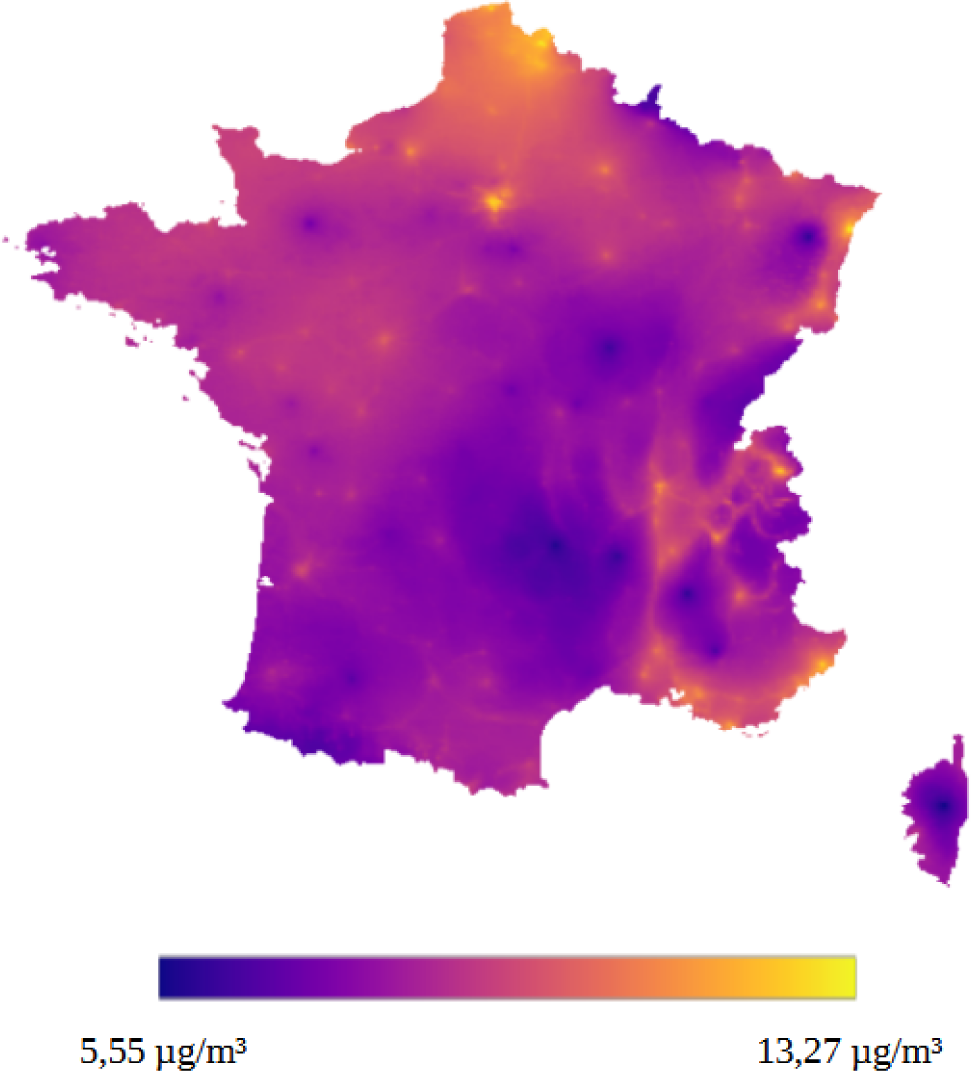
Spatial distribution of *PM*_2.5_ exposure in France Note: This figure maps the geographical distribution of the annual average exposure *PM*2.5 in 2019 on the French territory.

**Figure A3:**
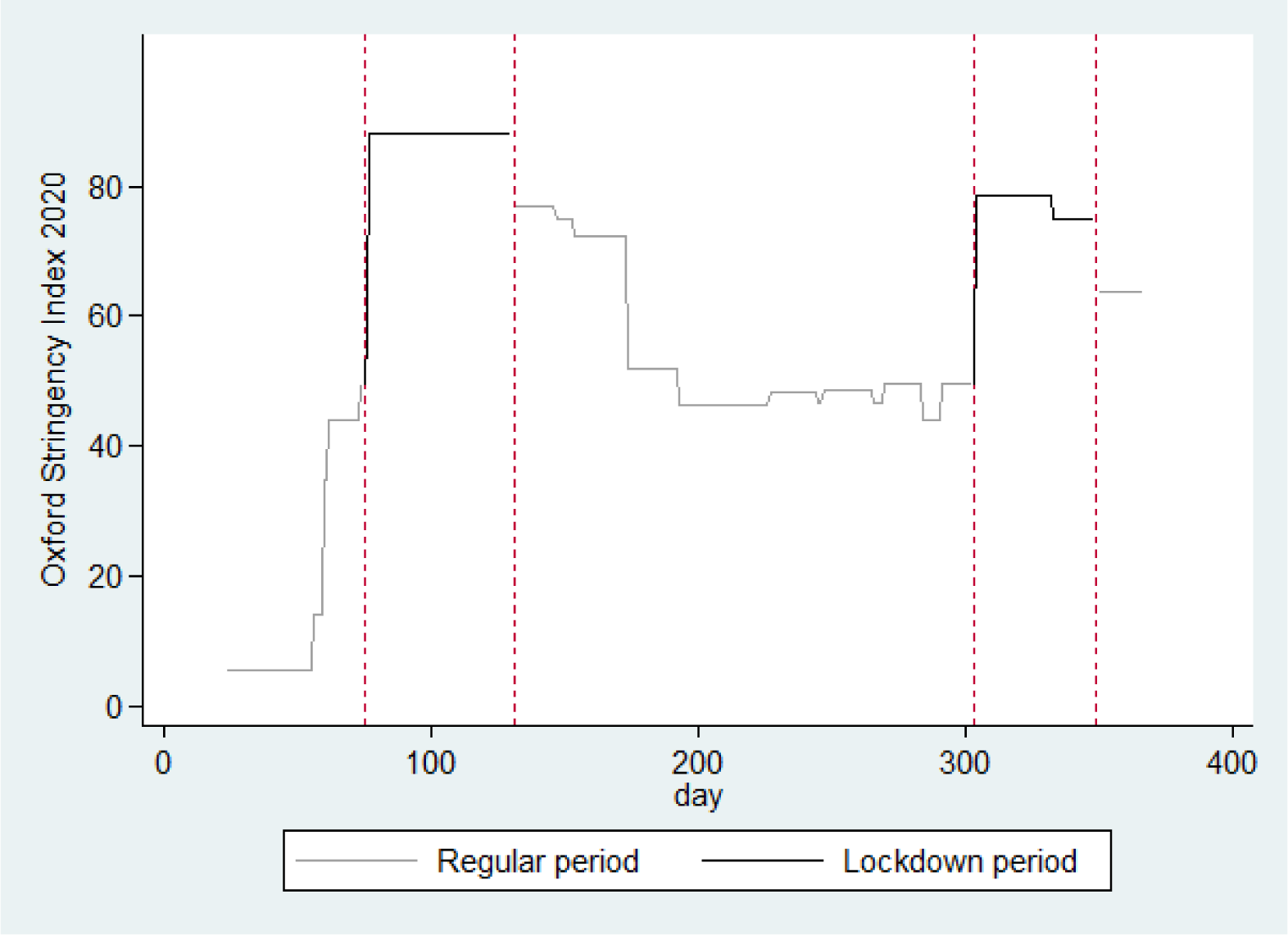
Evolution of the Oxford Stringency Index in France in 2020

## Appendix B Robustness checks

### B1 A regression discontinuity approach

One potential threat to the validity of our main identification assumption relates to the existence of shocks that would correlate with both air quality and lockdown policies, which could bias our main estimates. In particular, a recent literature highlighted a positive association between air pollution and the number of COVID-19 cases and related-mortality.^14^ Higher levels of air pollution could therefore indirectly induce governments to implement more stringent lockdown.

To further check the robustness of our main results, we estimate a Regression Discontinuity in Time (RDiT) model around lockdown dates. This alternative identification takes advantage of the fact that the implementation of lockdown policies represent a sudden and exogenous shock (Dang & Trinh, 2021). In this approach, observations just before the lockdown starting dates provide the counterfactual for observations immediately after the lockdown dates. It relies on the assumption that the exact starting date of the lockdown policy is randomized in the close neighborhood of the actual date.^15^

In practice, we estimate the following model using the rdrobust stata package (Calonico et al., 2017):

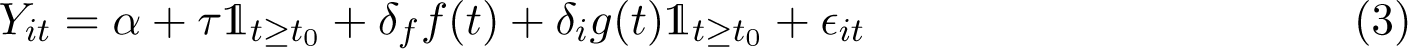

where *Y_it_*represents *PM*_2.5_ exposure in grid *i* in 2020 on day *t*, *t*_0_ indicates the cutoff (lockdown starting date) and 1*_t.≥.t_*_0_ is a dummy indicating lockdown implementation. *f* (.) and *g*(.) are flexible functions that we allow to differ on each side of the time cutoff. We cluster standard errors at the day level. We use the optimal data-driven bandwidth selection procedures proposed by Calonico et al. (2020).

Table B1 shows the result of the estimation of Equation (3) using different functional forms of the running variable: a linear model (column (1)), a quadratic model (column (2)) and a cubic model (column (3)). The first panel of the table shows the estimation for the first lockdown, which started on March, 17. The second panel of the table shows the estimation for the second lockdown, which started on October, 30. RDD results are consistent with the main results outlined in this paper, as a discontinuous increase in *PM*_2.5_ exposure is observed right after the implementation of each lockdown period. Figures 1(a) and 1(b), which respectively depicts the effects of the first and second lockdown estimated with the cubic model, further confirm the increase in *PM*_2.5_ exposure caused by restriction measures.

**Table B1:** Effects of lockdown on *PM*_2.5_ exposure in 2020: RDD estimations

| | (1)<br>$PM_{2.5}$ | (2)<br>$PM_{2.5}$ | (3)<br>$PM_{2.5}$ |
| --- | --- | --- | --- |
| *First lockdown* |  |  |  |
| RD_Estimate | 5.3573***<br>(1.1934) | 3.5473***<br>(.65153) | 3.3465***<br>(.69797) |
| Observations | 1236434 | 1236434 | 1236434 |
| *Second lockdown* |  |  |  |
| RD_Estimate | 1.8269**<br>(.93159) | .99007<br>(1.222) | 2.1329***<br>(.76227) |
| Observations | 1224741 | 1224741 | 1224741 |
| Model | Linear | Quadratic | Cubic |

Note:
This table shows the estimation of lockdown effects using a Regression Discontinuity in Time approach design. The bandwidth has been computed using the optimal data-driven procedure proposed by Calonico et al. \(2020\). Standard errors (in parentheses) are clustered at the day level. \* $p < 0.10$ , \*\* $p < 0.05$ , \*\*\* $p < 0.01$ .

**Figure B1:**
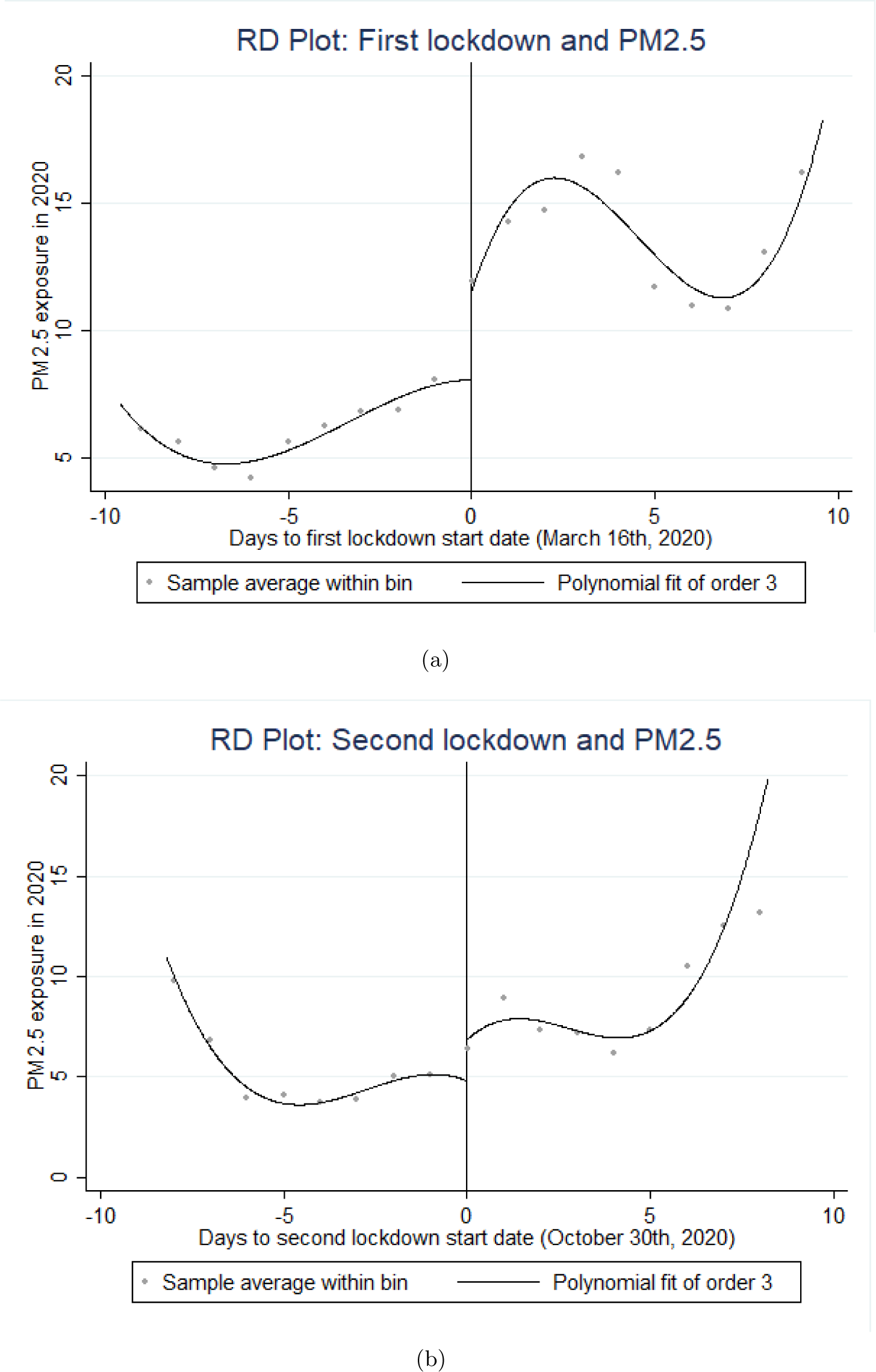
RD plot of lockdown effects

### B2 Weather controls

Our main econometric approach control for weather conditions in a very parsimonious way to avoid excessive computation time. In this appendix section, we provide evidence that controlling for weather conditions in a much more flexible way doesn’t change the nature of our main results. The high granularity and comprehensive scope of our data makes it possible to include a very large set of weather condition fixed effects. For each of our 4 weather variables (daily maximum temperatures, daily minimum temperatures, daily total precipitation and daily average wind speed), we generate indicators for the quartiles of these variables. We then generate a set of indicators for all possible interactions of these temperature (min and max), precipitation, wind speed and wind direction variables and include it in our main specification as *W_it_*. This regression model thus controls for more than 1,000 possible combinations of weather conditions. Table B2 reproduces the main results presented in Table 1 with a model including the large set of weather control variables, estimated on a 5% random subsample of the main sample. Those results reinforce the assumption that our estimates are not driven by unobserved meteorological factors that would be correlated with both lockdown implementation and *PM*_2.5_ concentration. Finally, our estimates are also robust to the omission of weather controls (Table B3).

**Table B2:** Effects of lockdown on *PM*_2.5_ exposure: large set of weather controls

|  | (1) | (2) | (3) | (4) |
| --- | --- | --- | --- | --- |
| | $PM_{2.5}$ 2020 | 2020 vs 2015-2019 | 2020 | 2020 vs 2015-2019 |
| Lockdown dummy | 2.4271***<br>(.3939) | 1.5201***<br>(.46194) |  |  |
| Oxford Stringency Index |  |  | .02108**<br>(.00871) | .03381***<br>(.00781) |
| Weather controls | ✓ | ✓ | ✓ | ✓ |
| Observations | 1410501 | 1409959 | 1319347 | 1318805 |

Standard errors in parentheses
\* $p < 0.10$ , \*\* $p < 0.05$ , \*\*\* $p < 0.01$

**Table B3:** Effects of lockdown on *PM*_2.5_ exposure: without weather controls

|  | (1) | (2) | (3) | (4) |
| --- | --- | --- | --- | --- |
| | $PM_{2.5}$ 2020 | 2020 vs 2015-2019 | 2020 | 2020 vs 2015-2019 |
| Lockdown dummy | 3.9309***<br>(.48366) | 2.559***<br>(.56552) |  |  |
| Oxford Stringency Index |  |  | .03899***<br>(.01169) | .05258***<br>(.01014) |
| Weather controls | . | . | . | . |
| Observations | 49211994 | 49211994 | 46119437 | 46119437 |

Standard errors in parentheses
\* $p < 0.10$ , \*\* $p < 0.05$ , \*\*\* $p < 0.01$

### B3 Explicit DiD

Our main econometric model can be viewed as an implicit difference-in-difference (DiD) design where the coefficient of interest is *β*_1_. It estimates the difference in excess pollution (time dimension) between days of lockdown (“treatment group”) and days of regular period (“comparison group‘”). This model identifies the causal effect of lockdown policies on excess pollution under the assumption that, absent lockdown, the evolution of pollution between 2015-2019 and 2020 would have been similar on average between days of lockdown and days of regular periods, after accounting for grid-specific seasonality and local weather conditions.

To further check the robustness of our results, we take advantage of the panel nature of our data to estimate an explicit DiD model, which includes day-of-the-year *×* grid fixed effects and uses *PM*_2.5_ exposure level as the main dependent variable. Formally, we estimate the following model, on a 5% random subsample of observations from our main sample:

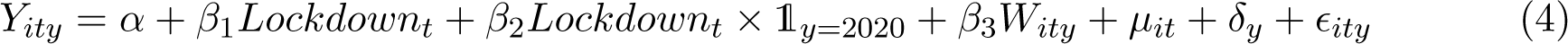

where *Y_ity_* represents *PM*_2.5_ exposure in grid *i* on day *t* of year *y* and *Lockdown_t_* is a dummy indicating lockdown periods. *µ_it_* represents the full set of day-of-the-year *×* grid fixed effects. *W_ity_*represents the set of weather control variables described in Section 2.2. Note that 1*_y_*_=2020_ is included in the set of year fixed effects *δ_y_* and that day and grid fixed effects are included in day *×* grid fixed effects *µ_it_*. We cluster all standard errors at the day level.

As shown in Table B4, the nature of the result is unchanged when implementing this explicit DiD model: on average, *PM*_2.5_ is 1.57 *µg/m*^3^ higher during days of lockdown.

**Table B4:** Effects of lockdown on *PM*_2.5_: Explicit DiD model

|  | (1) | (2) |
| --- | --- | --- |
| | $PM_{2.5}$ | $PM_{2.5}$ |
| Lockdown × 2020 | 1.5732***<br>(.36577) | |
| OSI × 2020 | | .02573***<br>(.00806) |
| Observations | 17190826 | 17036461 |

Standard errors in parentheses
\* $p < 0.10$ , \*\* $p < 0.05$ , \*\*\* $p < 0.01$

### B4 Permutation tests

We further check the robustness of our results to an alternative (randomization-based) inference approach by performing permutation tests. These tests randomly reassign the treatment status (i.e., lockdown) over days of the 2020 year. We then compare the observed difference in our outcome (*PM*_2.5_ exposure) between lockdown and regular days with the distribution of this difference in the placebo tests. This comparison allows us to assess whether the observed difference is likely to have been produced by chance - and not by the effects of lockdown policies. We implement these tests on a 5% random subsample of observations, using the ritest command (Heß, 2017) and simulating 100 permutations. As can be seen in Table B5, none of the 100 permutations performed provides a greater difference in our outcome than the one that is *actually* observed. This result further confirms that the likelihood that our estimated effect of lockdown policies is caused by chance is close to 0.

**Table B5:** Effects of lockdown on *PM*_2.5_: permutation tests

|  | (1) | (2) |
| --- | --- | --- |
| | $PM_{2.5}$ 2020 | $PM_{2.5}$ 2020 vs 2015-2019 |
| Lockdown | 2.51*** | 1.92*** |
|  | (0.01) | (0.01) |
|  | [0.00] | [0.00] |
| Observations | 2459583 | 2459583 |

Standard errors (in parentheses) clustered at the day level.
p-values (within brackets): \* $p < 0.10$ , \*\* $p < 0.05$ , \*\*\* $p < 0.01$

## Appendix C Spatial heterogeneity in the effects of lockdown

**Table C1:** Effects of lockdown on *PM*_2.5_: Heterogeneity by *PM*_2.5_ long-term exposure level

| | (1)<br>$PM_{2.5}$ 2020 | (2)<br>2020 vs 2015-2019 | (3)<br>2020 | (4)<br>2020 vs 2015-2019 |
| --- | --- | --- | --- | --- |
| Lockdown dummy | 2.5094***<br>(.4506) | 1.8969***<br>(.49141) |  |  |
| Oxford Stringency Index (OSI) |  |  | .02056**<br>(.00999) | .03911***<br>(.00845) |
| $PM_{2.5}$ exposure 2009-2019 | | -.44317***<br>(.04764) | | -.89601***<br>(.15975) |
| $PM_{2.5}$ exposure 2009-2019 × Lockdown | .43694***<br>(.10504) | .24796**<br>(.12552) | | |
| $PM_{2.5}$ exposure 2009-2019 × OSI | | | .0062**<br>(.00251) | .00899***<br>(.00266) |
| Observations | 49211628 | 49211628 | 46119094 | 46119094 |

Standard errors in parentheses
\* $p < 0.10$ , \*\* $p < 0.05$ , \*\*\* $p < 0.01$

**Table C2:** Effects of lockdown on *PM*_2.5_: Heterogeneity by *NO*_2_ long-term exposure level

| | (1)<br>$PM_{2.5}$ 2020 | (2)<br>2020 vs 2015-2019 | (3)<br>2020 | (4)<br>2020 vs 2015-2019 |
| --- | --- | --- | --- | --- |
| Lockdown dummy | 1.4667**<br>(.58172) | .79774<br>(.71047) |  |  |
| Oxford Stringency Index (OSI) |  |  | -.0015<br>(.01132) | .00032<br>(.01204) |
| $NO_2$ exposure 2009-2019 | | -.14905***<br>(.01651) | | -.30326***<br>(.05116) |
| $NO_2$ exposure 2009-2019 × Lockdown | .07926**<br>(.03691) | .0834**<br>(.04176) | | |
| $NO_2$ exposure 2009-2019 × OSI | | | .00168**<br>(.00083) | .00294***<br>(.00086) |
| Observations | 49211628 | 49211628 | 46119094 | 46119094 |

Standard errors in parentheses
\* $p < 0.10$ , \*\* $p < 0.05$ , \*\*\* $p < 0.01$

**Figure C1:**
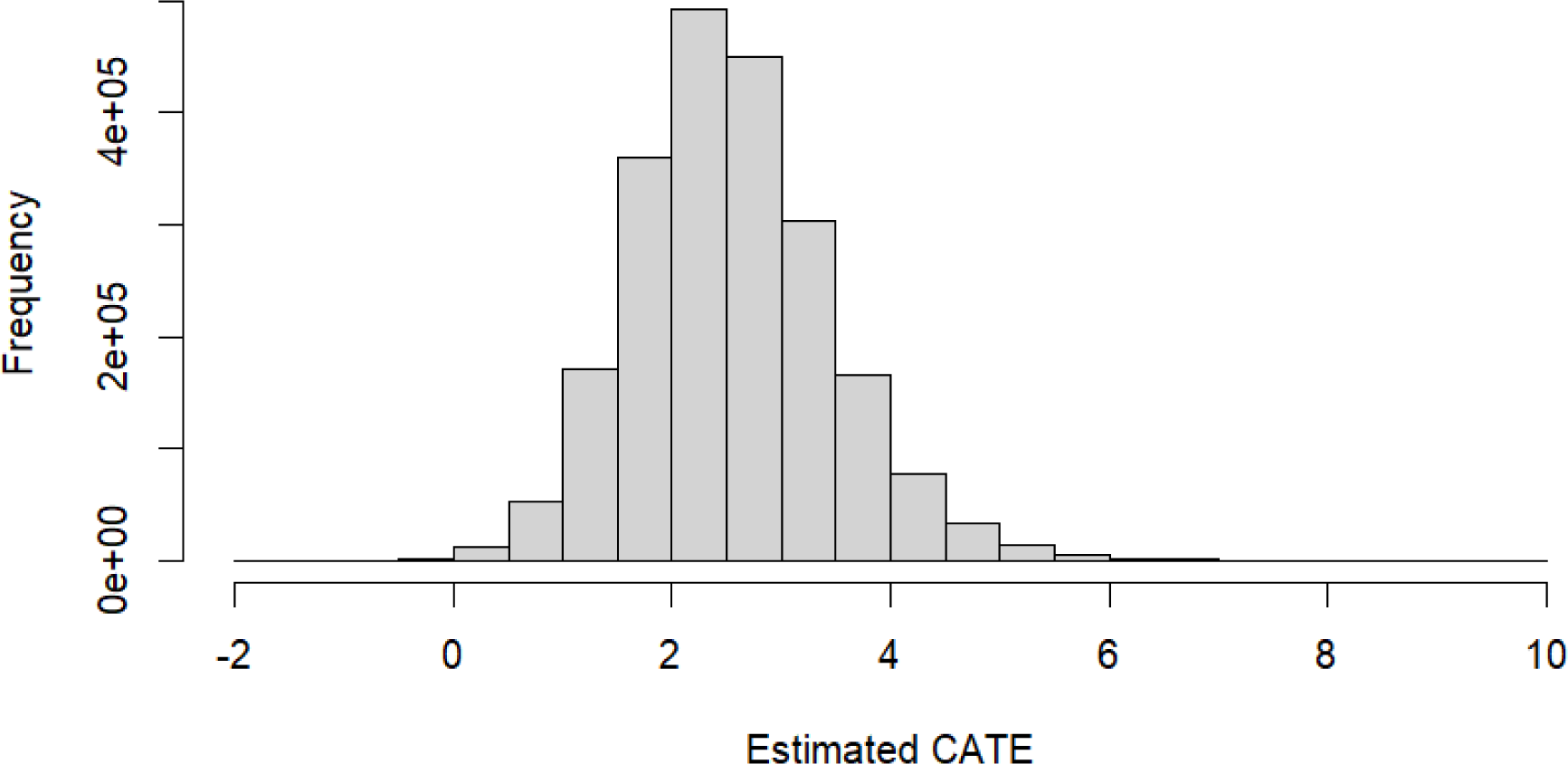
Conditional Average Treatment Effects distribution Note: This figure shows the distribution of the conditional average treatment effects estimated from the GRF procedure.

**Figure C2:**
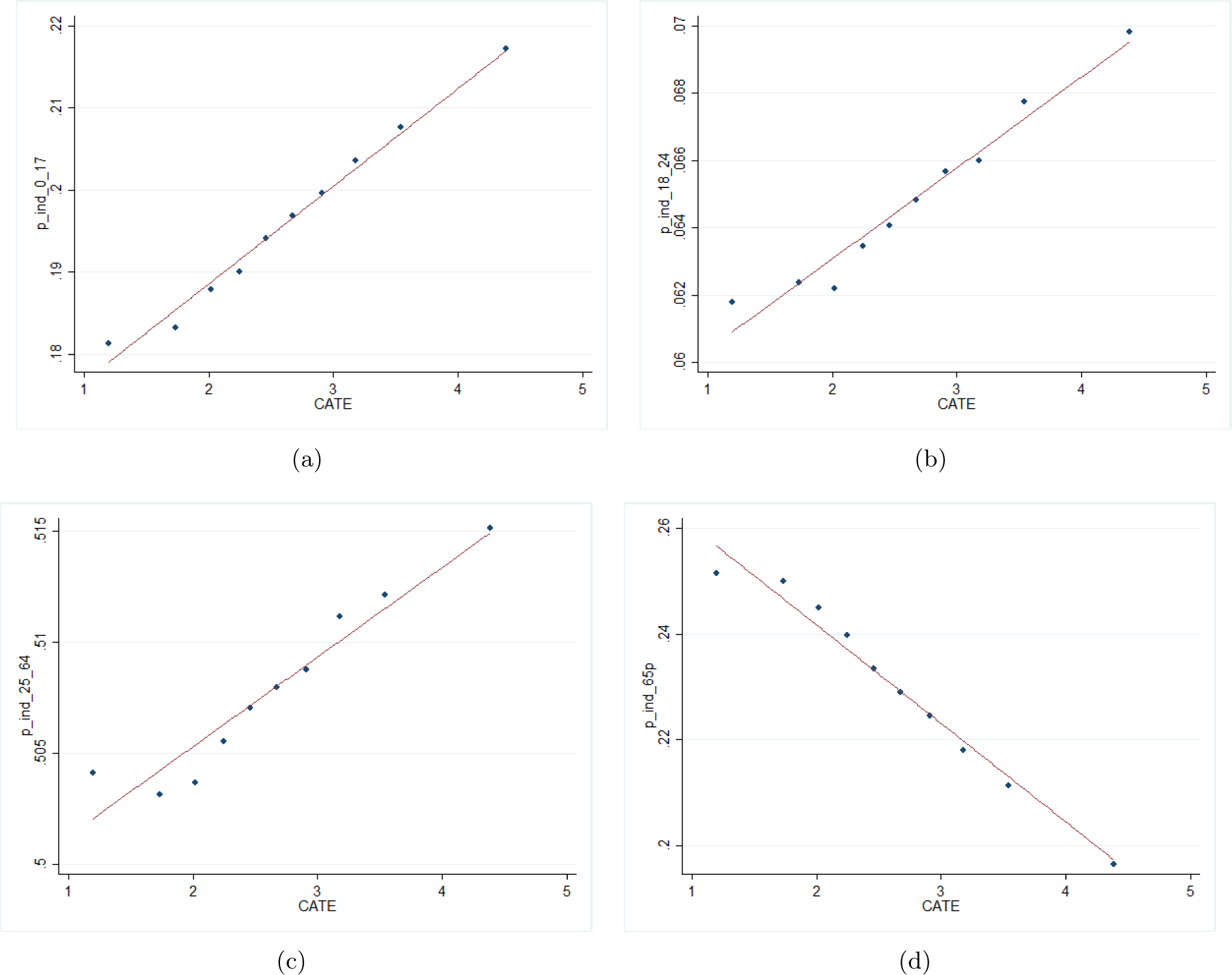
Conditional Average Treatment Effects and Inhabitants Age *.N..o..t..e..:.* Figures C2 (a) to (d) respectively show the distribution of grid cells average proportion of inhabitants between 0 and 17 years old, between 18 and 24 years old, between 25 and 64 years old, and above 65 years old, by deciles of the conditional average treatment effects estimated from the GRF procedure.

**Figure C3:**
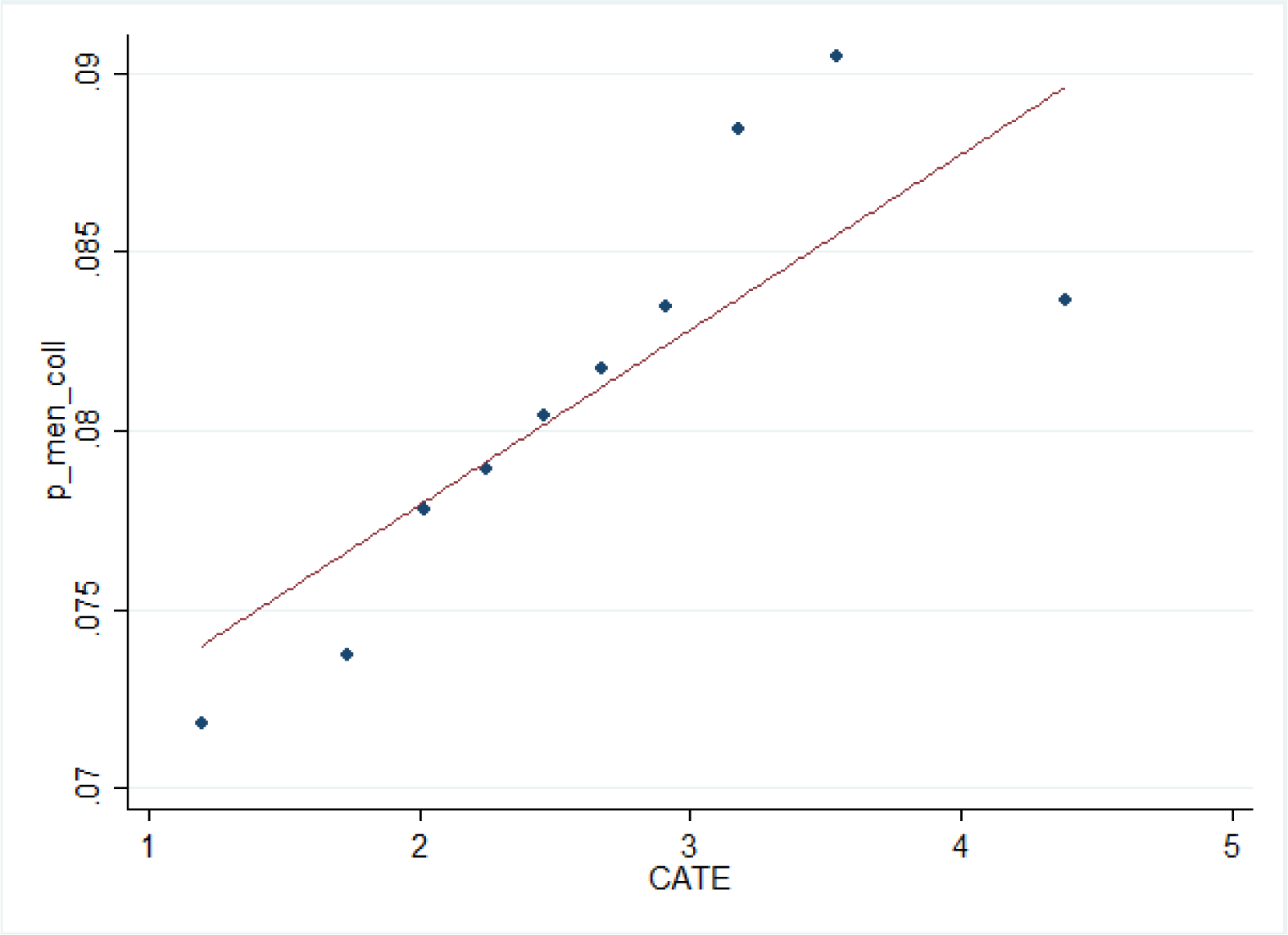
Conditional Average Treatment Effects and share of Households in Collective Dwellings Note: This figure shows the distribution of grid cells’ average share of households living in collective dwellings by deciles of the conditional average treatment effects estimated from the GRF procedure.

**Figure C4:**
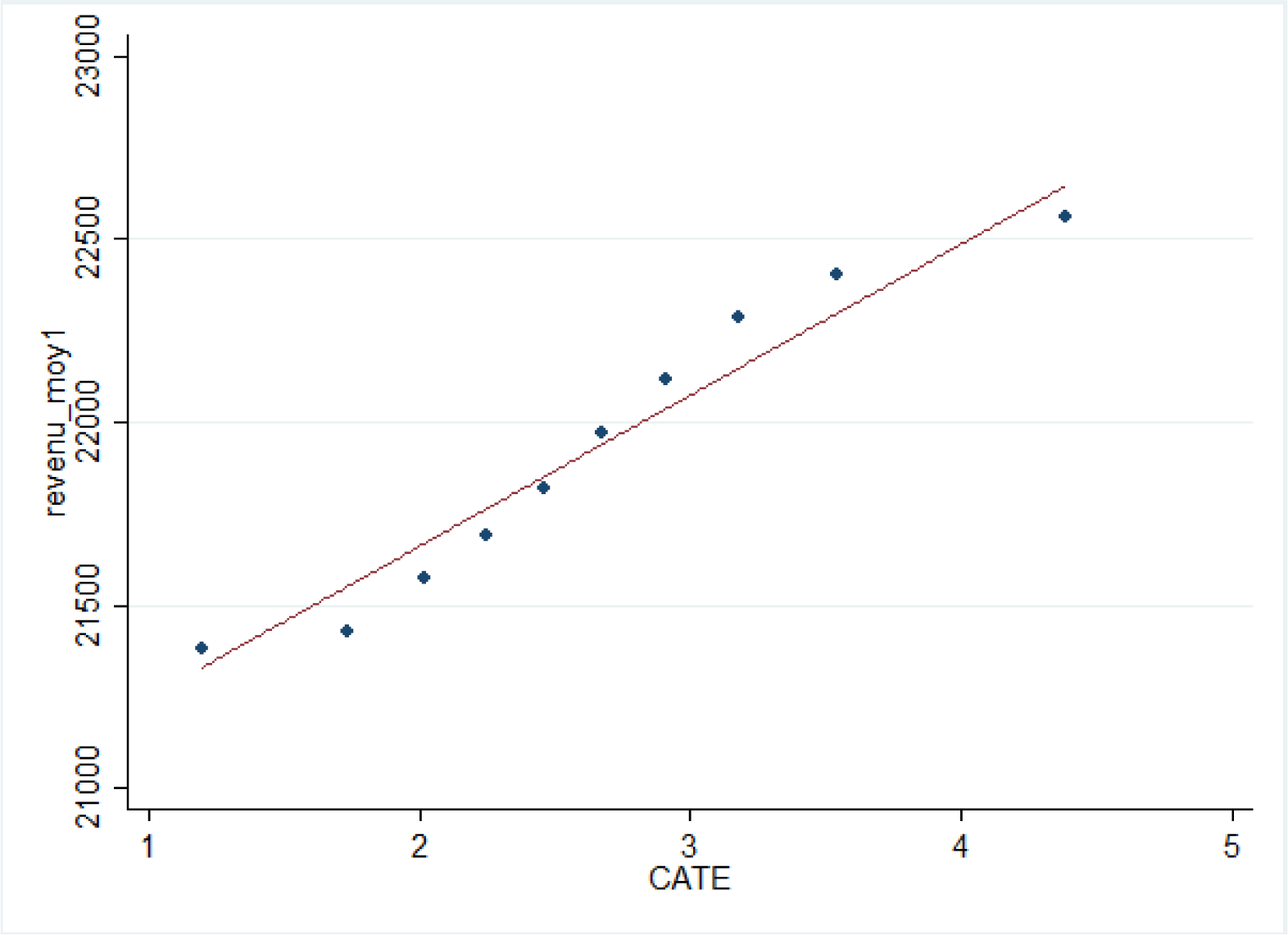
Conditional Average Treatment Effects and Household Income Note: This figure shows the distribution of grid cells’ average household income by deciles of the conditional average treatment effects estimated from the GRF procedure.

## Footnotes

1 By contrast, we do not exploit directly measures based on local station data (industrial or traffic), whose purpose is to measure pollution emissions - rather than exposure - at a local level. These monitoring stations are sparse and strategically placed to capture locally produced emissions (e.g., from road traffic or industrial activities).

2 This literature highlights in particular the negative role played by fine particulate matter (Pope et al., 2002; Lepeule et al., 2012; Lelieveld et al., 2015; Ciarelli et al., 2019; Deryugina et al., 2019). For example, the Environmental Protection Agency (EPA) estimates that *PM*2.5 is responsible for over 90% of air pollution-related health damages in the US (US Environmental Protection Agency, 2011). A growing body of evidence also documents the adverse economic effects of air pollution on non-health outcomes, including labor supply, cognitive performance and labor productivity, educational attainment or crime. See Aguilar-Gomez et al. (2022) for a recent review of this evidence.

3 Average exposure to *NO*2 in France in 2020 is 8.3 *µg/m*^3^.

4 In a critical review of more than 200 papers, Gkatzelis et al. (2021) highlight the significant effects of meteorological conditions on pollutant concentrations. Barŕe et al. (2021) further show the importance of accounting for weather conditions to estimate *NO*2 changes induced by lockdown policies.

5 There are 875 weather stations in our dataset. The average distance between a grid cell from our pollution data and the closest weather station is 11.8km. The maximum distance is 50 km, but more than 99% of grid cells are located less than 27kms away from the closest weather station.

6 Overall in 2020, there was 49,495 deaths in excess in French urban areas in comparison with the average of 2018 and 2019 (Brandily et al., 2021).

7 Analyses based on a formal Regression Discontinuity in Time (RDiT) approach confirm the existence of a significant discontinuity in *PM*2.5 at both time cutoffs (see Appendix B1).

8 For computational reasons, our main estimations include a continuous specification of our 4 weather variables (total precipitations, surface maximum temperatures, surface minimum temperatures, wind speed) as well as a dummy for each of the four possible wind directions. In Appendix B2, we provide evidence that our results are unchanged when including a very large set of weather fixed effects, based on bins for each of the four variables and their interactions.

9 Note that day-of-the-year fixed effects and grid fixed effects are included in *κit*.

10 In Appendix B3, we show that our results are robust to estimating a formal (explicit) DiD model. [11 The analyses conducted in this section use the R package grf, version 2.0.2 (Tibshirani et al. (2021)).] [12 Conversely, when *H*0 is not rejected, this does not necessarily imply that there is no significant heterogeneity in treatment effects, it may also mean that the causal forest procedure does not produce relevant predictions of treatment effects.] [13 These two variables respectively account for 72% and 16% of the splits. Other variables that appear in at least 2-3% of the splits relate to the age distribution of inhabitants, to their average income and to the proportion of households living in collective dwellings.] [14 See for example Coker et al. (2020); Wu et al. (2020); Isphording & Pestel (2021); Persico & Johnson (2021); Weaver et al. (2022) or Becchetti et al. (2022).] [15 RDiT methods have been widely implemented in the economic literature that estimate the causal impact of specific shocks on air quality (Davis, 2008; Auffhammer & Kellogg, 2011; Chen & Whalley, 2012; Gallego et al., 2013; Dang & Trinh, 2021). However, these methods may suffer from methodological limitations, especially in the presence of time varying treatment effects or autoregression (Hausman & Rapson, 2018).]

## Notes

### Competing Interest Statement

The authors have declared no competing interest.

### Funding Statement

This study was funded by the ADEME grant AQACIA 2162D0019 - Air-COV project.

## References

1. Adam, M. G., Tran, P. T., & Balasubramanian, R. 2021. “Air quality changes in cities during the COVID-19 lockdown: A critical review”. Atmospheric Research, 264:105823.

2. Aguilar-Gomez, S., Dwyer, H., Graff Zivin, J., & Neidell, M. 2022. “This is air: The “nonhealth” effects of air pollution”. Annual Review of Resource Economics, 14:403–425.

3. Äıchi, L. & Husson, J. 2015. “Pollution de l’air: le coût de l’inaction”. Rapport pour la commission d’enqûete sur le coût économique et financier de la pollution de l’air pour le śenat, (610):1–14.

4. Allcott, H., Braghieri, L., Eichmeyer, S., & Gentzkow, M. 2020. “The welfare effects of social media”. American Economic Review, 110(3):629–676.

5. Athey, S. & Wager, S. 2019. “Estimating treatment effects with causal forests: An application”. Observational Studies, 5(2):37–51.

6. Athey, S., Tibshirani, J., & Wager, S. 2019. “Generalized random forests”. The Annals of Statistics, 47(2):1148–1178.

7. Auffhammer, M. & Kellogg, R. 2011. “Clearing the air? The effects of gasoline content regulation on air quality”. American Economic Review, 101(6):2687–2722.

8. Barŕe, J., Petetin, H., Colette, A., Guevara, M., Peuch, V.-H., Rouil, L., Engelen, R., Inness, A., Flemming, J., Garćıa-Pando, C. P., et al. 2021. “Estimating lockdown-induced European NO2 changes using satellite and surface observations and air quality models”. Atmospheric chemistry and physics, 21(9):7373–7394.

9. Bartoňová, A., Colette, A., Zhang, H., Fons, J., Liu, H.-Y., Brzezina, J., Chantreux, A., Couvidat, F., Guerreiro, C., Guevara, M., et al. 2022. “The Covid-19 pandemic and environmental stressors in Europe: synergies and interplays”. ETC/ATNI Report Kjeller.

10. Becchetti, L., Beccari, G., Conzo, G., Conzo, P., De Santis, D., & Salustri, F. 2022. “Particulate matter and COVID-19 excess deaths: Decomposing long-term exposure and short-term effects”. Ecological Economics, 194:107340.

11. Berman, J. D. & Ebisu, K. 2020. “Changes in US air pollution during the COVID-19 pandemic”. Science of the total environment, 739:139864.

12. Brandily, P., Bŕebion, C., Briole, S., & Khoury, L. 2021. “A poorly understood disease? The impact of COVID-19 on the income gradient in mortality over the course of the pandemic”. European Economic Review, 140:103923.

13. Briole, S., Gurgand, M., Maurin, É., McNally, S., Ruiz-Valenzuela, J., & Sańın, D. 2022. “The Making of Civic Virtues: A School-Based Experiment in Three Countries”. IZA DP No. 15141.

14. Brodeur, A., Cook, N., & Wright, T. 2021. “On the effects of COVID-19 safer-at-home policies on social distancing, car crashes and pollution”. Journal of Environmental Economics and Management, 106:102427.

15. Calonico, S., Cattaneo, M. D., Farrell, M. H., & Titiunik, R. 2017. “rdrobust: Software for regression-discontinuity designs”. The Stata Journal, 17(2):372–404.

16. Calonico, S., Cattaneo, M. D., & Farrell, M. H. 2020. “Optimal bandwidth choice for robust bias-corrected inference in regression discontinuity designs”. The Econometrics Journal, 23(2): 192–210.

17. Carter, M. R., Tjernström, E., & Toledo, P. 2019. “Heterogeneous impact dynamics of a rural business development program in Nicaragua”. Journal of Development Economics, 138:77–98.

18. Chen, Y. & Whalley, A. 2012. “Green infrastructure: The effects of urban rail transit on air quality”. American Economic Journal: Economic Policy, 4(1):58–97.

19. Chernozhukov, V., Demirer, M., Duflo, E., & Fernandez-Val, I. 2018. “Generic Machine Learning Inference on Heterogeneous Treatment Effects in Randomized Experiments, with an Application to Immunization in India”. National Bureau of Economic Research.

20. Ciarelli, G., Colette, A., Schucht, S., Beekmann, M., Andersson, C., Manders-Groot, A., Mircea, M., Tsyro, S., Fagerli, H., Ortiz, A. G., et al. 2019. “Long-term health impact assessment of total PM2. 5 in Europe during the 1990–2015 period”. Atmospheric Environment: X, 3:100032.

21. Citepa. 2022. “Inventaire des émissions de polluants atmosphériques et de gaz à effet de serre en France – Format Secten”.

22. Coker, E. S., Cavalli, L., Fabrizi, E., Guastella, G., Lippo, E., Parisi, M. L., Pontarollo, N., Rizzati, M., Varacca, A., & Vergalli, S. 2020. “The effects of air pollution on COVID-19 related mortality in northern Italy”. Environmental and Resource Economics, 76:611–634.

23. Currie, J. & Walker, R. 2019. “What do economists have to say about the Clean Air Act 50 years after the establishment of the Environmental Protection Agency?”. Journal of Economic Perspectives, 33(4):3–26.

24. Currie, J., Voorheis, J., & Walker, R. 2023. “What caused racial disparities in particulate exposure to fall? New evidence from the Clean Air Act and satellite-based measures of air quality”. American Economic Review, 113(1):71–97.

25. Daellenbach, K. R., Uzu, G., Jiang, J., Cassagnes, L.-E., Leni, Z., Vlachou, A., Stefenelli, G., Canonaco, F., Weber, S., Segers, A., et al. 2020. “Sources of particulate-matter air pollution and its oxidative potential in Europe”. Nature, 587(7834):414–419.

26. Dang, H.-A. H. & Trinh, T.-A. 2021. “Does the COVID-19 lockdown improve global air quality? New cross-national evidence on its unintended consequences”. Journal of Environmental Economics and Management, 105:102401.

27. Davis, L. W. 2008. “The effect of driving restrictions on air quality in Mexico City”. Journal of Political Economy, 116(1):38–81.

28. Deryugina, T., Heutel, G., Miller, N. H., Molitor, D., & Reif, J. 2019. “The mortality and medical costs of air pollution: Evidence from changes in wind direction”. American Economic Review, 109(12):4178–4219.

29. Faridi, S., Yousefian, F., Janjani, H., Niazi, S., Azimi, F., Naddafi, K., & Hassanvand, M. S. 2021. “The effect of COVID-19 pandemic on human mobility and ambient air quality around the world: A systematic review”. Urban Climate, 38:100888.

30. Gallego, F., Montero, J.-P., & Salas, C. 2013. “The effect of transport policies on car use: Evidence from Latin American cities”. Journal of Public Economics, 107:47–62.

31. Gkatzelis, G. I., Gilman, J. B., Brown, S. S., Eskes, H., Gomes, A. R., Lange, A. C., McDonald, B. C., Peischl, J., Petzold, A., Thompson, C. R., et al. 2021. “The global impacts of covid-19 lockdowns on urban air pollutiona critical review and recommendations”. Elementa: Science of the Anthropocene, 9(1).

32. Haaland, I. & Roth, C. 2020. “Labor market concerns and support for immigration”. Journal of Public Economics, 191:104256.

33. Hale, T., Angrist, N., Goldszmidt, R., Kira, B., Petherick, A., Phillips, T., Webster, S., Cameron-Blake, E., Hallas, L., Majumdar, S., et al. 2021. “A global panel database of pandemic policies (Oxford COVID-19 Government Response Tracker)”. Nature human behaviour, 5(4):529–538.

34. Hausman, C. & Rapson, D. S. 2018. “Regression discontinuity in time: Considerations for empirical applications”. Annual Review of Resource Economics, 10:533–552.

35. Heß, S. 2017. “Randomization inference with Stata: A guide and software”. The Stata Journal, 17 (3):630–651.

36. Isphording, I. E. & Pestel, N. 2021. “Pandemic meets pollution: poor air quality increases deaths by COVID-19”. Journal of Environmental Economics and Management, 108:102448.

37. Lavaine, E., Majerus, P., & Treich, N. 2020. “Health, air pollution, and animal agriculture”. *Review of Agricultural*, Food and Environmental Studies, 101:517–528.

38. Lelieveld, J., Evans, J. S., Fnais, M., Giannadaki, D., & Pozzer, A. 2015. “The contribution of outdoor air pollution sources to premature mortality on a global scale”. Nature, 525(7569): 367–371.

39. Lepeule, J., Laden, F., Dockery, D., & Schwartz, J. 2012. “Chronic exposure to fine particles and mortality: an extended follow-up of the Harvard Six Cities study from 1974 to 2009”. Environmental health perspectives, 120(7):965–970.

40. Mahato, S., Pal, S., & Ghosh, K. G. 2020. “Effect of lockdown amid COVID-19 pandemic on air quality of the megacity Delhi, India”. Science of the total environment, 730:139086.

41. Menut, L., Bessagnet, B., Khvorostyanov, D., Beekmann, M., Blond, N., Colette, A., Coll, I., Curci, G., Foret, G., Hodzic, A., et al. 2013. “CHIMERE 2013: a model for regional atmospheric composition modelling”. Geoscientific model development, 6(4):981–1028.

42. Persico, C. L. & Johnson, K. R. 2021. “The effects of increased pollution on COVID-19 cases and deaths”. Journal of Environmental Economics and Management, 107:102431.

43. Pope, C. A., Burnett, R. T., Thun, M. J., Calle, E. E., Krewski, D., Ito, K., & Thurston, G. D. 2002. “Lung cancer, cardiopulmonary mortality, and long-term exposure to fine particulate air pollution”. Jama, 287(9):1132–1141.

44. Salesse, C. 2022. “Inequality in exposure to air pollution in France: bringing pollutant cocktails into the picture”. CEE-M Working Paper 2022–11.

45. Schneider, R., Masselot, P., Vicedo-Cabrera, A. M., Sera, F., Blangiardo, M., Forlani, C., Douros, J., Jorba, O., Adani, M., Kouznetsov, R., et al. 2022. “Differential impact of government lockdown policies on reducing air pollution levels and related mortality in Europe”. Scientific reports, 12 (1):726.

46. Sicard, P., De Marco, A., Agathokleous, E., Feng, Z., Xu, X., Paoletti, E., Rodriguez, J. J. D., & Calatayud, V. 2020. “Amplified ozone pollution in cities during the COVID-19 lockdown”. Science of the Total Environment, 735:139542.

47. Sicard, P., Agathokleous, E., De Marco, A., Paoletti, E., & Calatayud, V. 2021. “Urban population exposure to air pollution in Europe over the last decades”. Environmental Sciences Europe, 33 (1):1–12.

48. Sylvia, S., Warrinnier, N., Luo, R., Yue, A., Attanasio, O., Medina, A., & Rozelle, S. 2021. “From quantity to quality: Delivering a home-based parenting intervention through China’s family planning cadres”. The Economic Journal, 131(635):1365–1400.

49. Tibshirani, J., Athey, S., Friedberg, R., Hadad, V., Hirshberg, D., Miner, L., Sverdrup, E., Wager, S., & Wright, M. “Grf: Generalized Random Forests (version 2.0.2). R”, 2021.

50. US Environmental Protection Agency. “The benefits and costs of the Clean Air Act from 1990 to 2020”, 2011.

51. Venter, Z. S., Aunan, K., Chowdhury, S., & Lelieveld, J. 2020. “COVID-19 lockdowns cause global air pollution declines”. Proceedings of the National Academy of Sciences, 117(32):18984–18990.

52. Weaver, A. K., Head, J. R., Gould, C. F., Carlton, E. J., & Remais, J. V. 2022. “Environmental factors influencing COVID-19 incidence and severity”. Annual review of public health, 43:271–291.

53. WHO. 2021. WHO global air quality guidelines: particulate matter (PM2. 5 and PM10), ozone, nitrogen dioxide, sulfur dioxide and carbon monoxide. World Health Organization.

54. Wu, X., Nethery, R. C., Sabath, M. B., Braun, D., & Dominici, F. 2020. “Air pollution and COVID-19 mortality in the United States: Strengths and limitations of an ecological regression analysis”. Science advances, 6(45):eabd4049.

